# Food insecurity and the risk of HIV acquisition: Findings from population-based surveys in six sub-Saharan African countries (2016-2017)

**DOI:** 10.1101/2021.09.27.21263917

**Authors:** Andrea Low, Elizabeth Gummerson, Amee Schwitters, Rogerio Bonifacio, Mekleet Teferi, Nicholus Mutenda, Sarah Ayton, James Juma, Claudia Ahpoe, Choice Ginindza, Hetal Patel, Sam Biraro, Karam Sachathep, Avi Hakim, Danielle T. Barradas, Ahmed Saadani Hassani, Wilford Kirungi, Keisha Jackson, Leah H. Goeke, Neena M. Philip, Lloyd Mulenga, Jennifer Ward, Steven Hong, George Rutherford, Sally Findley

## Abstract

**Introduction:** Food insecurity has a bidirectional relationship with HIV infection, with hunger driving compensatory risk behaviors, while infection can increase poverty. We used a laboratory recency assay to estimate the timing of HIV infection vis-à-vis the timing of severe food insecurity (SFI).

**Methods:** Data from population-based surveys in Zambia, Eswatini, Lesotho, Uganda, and Tanzania and Namibia were used. We defined SFI as having no food ≥three times in the past month. Recent HIV infection was identified using the HIV-1 LAg avidity assay, with a viral load (>1000 copies/ml) and no detectable antiretrovirals indicating an infection in the past 6 months. Logistic regression was conducted to assess correlates of SFI. Poisson regression was conducted on pooled data, adjusted by country to determine the association of SFI with recent HIV infection and risk behaviors, with effect heterogeneity evaluated for each country. All analyses were done using weighted data.

**Results:** Of 112,955 participants aged 15-59, 10.3% lived in households reporting SFI. SFI was most common in urban, woman-headed households. Among women and not men, SFI was associated with a two-fold increase in risk of recent HIV infection (adjusted relative risk [aRR] 2.08, 95% CI 1.09-3.97), with lower risk in high prevalence countries (Eswatini and Lesotho). SFI was associated with transactional sex (aRR 1.28, 95% CI 1.17-1.41), a history of forced sex (aRR 1.36, 95% CI 1.11-1.66), and condom-less sex with a partner of unknown or positive HIV status (aRR 1.08, 95% CI 1.02-1.14) in all women, and intergenerational sex (partner ≥10 years older) in women aged 15-24 (aRR 1.23, 95% CI 1.03-1.46), although this was heterogeneous. Recent receipt of food support was protective (aRR 0.36, 95% CI 0.14-0.88).

**Conclusion:** SFI increased risk for HIV acquisition in women by two-fold. Worsening food scarcity due to climactic extremes could imperil HIV epidemic control.

**SUMMARY:** *What is already known:*

- The link between food insecurity and the adoption of high-risk sexual behaviors as a coping mechanism has been shown in several settings.
- HIV infection can also drive food insecurity due to debilitating illness reducing productivity, the costs of treatment diverting money from supplies, and potentially reduced labor migration.
- Food insecurity has been associated with chronic HIV infection, but it has not been linked with HIV acquisition.

*What are the new findings:*

- This study of 112,955 adults across six countries in sub-Saharan Africa provides unique information on the association between acute food insecurity and recent HIV infection in women, as well as the potential behavioral and biological mediators, including community viremia as a measure of infectiousness.
- The data enabled a comprehensive analysis of factors associated with risk of infection, and how these factors differed by country and gender. Women living in food insecure households had a two-fold higher risk of recent HIV acquisition, and reported higher rates of transactional sex, early sexual debut, forced sex, intergenerational sex and sex without a condom with someone of unknown or positive HIV status. This pattern was not seen in men.
- This study is also the first to demonstrate a protective association for food support, which was associated with a lower risk of recent HIV infection in women.

*What do the new findings imply:*

- In light of worsening food insecurity due to climate change and the recent COVID-19 pandemic, our results support further exploration of gender-specific pathways of response to acute food insecurity, particularly how women’s changes in sexual behavior heighten their risk of HIV acquisition.
- These and other data support the inclusion of food insecurity in HIV risk assessments for women, as well as the exploration of provision of food support to those households at highest risk based on geographic and individual factors.

## INTRODUCTION

Climate change and its consequences are having a profound and escalating impact on global health. Acute events such as cyclones and flooding are predicted to become more frequent and severe, as are slower-onset changes such as drought and temperature extremes. These changes impact all domains of food security, including availability, access and utilization.^1-3^ Trends in world hunger have slowly reverted from a steady decline to a yearly increase, with a particular rise in sub-Saharan Africa (SSA), where almost 20 percent of the population is undernourished.^4^ Even predating the COVID-19 pandemic, models predicted that the risk of hunger and malnutrition globally could increase by 20% by 2050, generating humanitarian need in 200 million people per year,^5^ with the problem currently exacerbated by the economic impacts of the pandemic.^6 7^ Food insecurity impacts every facet of society, including political stability, economic productivity, and population displacement.

Food insecurity can be either acute or chronic.^8^ The primary drivers of transitory food insecurity relate to prices and availability, which are sensitive to environmental stressors, whereas chronic food insecurity is driven more by poverty.^9^ Acute food insecurity is a sensitive measure of economic shock and can capture changes in wealth that might prompt changes in health-related behaviors or trigger coping strategies, such as exchanging sex for food.^10-12^

The HIV pandemic has had a bidirectional link to food insecurity,^13^ as the associated health consequences can drive lower productivity and decreased labor mobility, whereas food insecurity can increase HIV risk behaviors, disruptions in care and higher mortality.^11-13^ The impact has been assumed to be gendered in that women are particularly vulnerable to income shocks and to disruption to access to health resources.^14^ Food insecurity has also been associated with lower efficacy of antiretroviral treatment (ART) due to drug malabsorption or decreased adherence, with virologic failure.^15^ As countries pursue the new UNAIDS 95-95-95 goals, weather extremes disrupting food production and supplies could jeopardize epidemic control, both in terms of increased risk behaviors, as well as disruption of treatment due to displacement or poverty, impacting access to testing and ART services. This results in increasing community-level infectiousness, driving the synergistic relationship between land degradation, vulnerability to drought, food insecurity and HIV transmission.^16 17^

The Population-Based HIV Impact Assessments (PHIAs), a series of national household-based surveys which collected data on the prevalence of HIV, recent HIV infection and viral load suppression (VLS), were conducted in several countries in SSA beginning in 2015. These surveys provide a unique opportunity to assess the relationship between food insecurity and HIV incidence in a large representative cohort of individuals. We used a theoretical framework to explore the relationships between food shortages, HIV and behavioral and biological mediators [(Supplementary Figure 1, appendix p 2)].

## METHODS

### Survey Design

We used data from all PHIA surveys collecting data on household food availability before 2018 (Eswatini, Lesotho, Namibia, Tanzania, Uganda, and Zambia). Surveys employed a two-stage sampling design to select a nationally representative sample of people aged 0-59 years or greater in each country.^18^ Consenting heads of households provided a roster of household members, who separately consented to interviews and household-based HIV testing. A guardian or parent provided permission for adolescent minors who were then asked for assent for all procedures. Written or verbal (Tanzania and Uganda) informed consent/assent was documented via electronic signature, with witnesses verifying consent for illiterate individuals. The PHIA protocol and data collection tools were approved by national ethics committees for each country, and the institutional review boards at Columbia University Irving Medical Center, the US Centers for Disease Control and Prevention (CDC) and the University of California, San Francisco in the case of Namibia. The period during which the surveys were conducted spanned different climate contexts, ranging from intense drought to overly wet conditions and flooding, described in the appendix [(Supplementary Figures 2-5, appendix pp 3-5)].

### Patient and public involvement

Patients were not involved directly in the formation of this study, although representatives from organizations representing people living with HIV were consulted as part of the questionnaire design, and as part of dissemination activities.

### Procedures

Interviewers administered the household questionnaire, which captured data from the household head on household assets, receipt of social support in the past three months, and access to food as measured by the Household Food Insecurity Access Scale Indicator Guide. We defined severe food insecurity as a household having no food in the house at least three times in the past four weeks. Receipt of food support was defined as having received food regardless of receipt of other support. The Dependent Ratio was calculated by dividing the number of children on the household roster by the number of adults, multiplied by 100, and then divided into quartiles. The adult questionnaire was administered to all eligible participants aged 15 and older during face-to-face interviews using Google Nexus 9 tablets. The questionnaire included questions on lifetime (excluding Tanzania) and recent sexual behaviors (past 12 months), and on characteristics of the three most recent sexual partners. Transactional sex was defined as having exchanged sex for material support or having sold sex in the past 12 months. Early sexual debut was sex occurring before age 15, and intergenerational sex as partnering with someone at least 10 years older. High-risk sex was defined as having sex without a condom with someone with an unknown or positive HIV status. Sampling design and questionnaire specifics are included in the appendix [(pp 6-7)].

Survey staff tested participants for HIV using the national algorithm. HIV RNA in plasma and dried blood spots (DBS) was measured using real-time PCR. Laboratory staff at the University of Cape Town conducted qualitative screening for detection of the most commonly used antiretrovirals (ARVs) with long half-lives on DBS specimens from all HIV-infected adults. Staff used the HIV-1 limited antigen (LAg) avidity immunoassay to classify recent infection in HIV-positive samples, where samples with a normalized optical density below 1.5 that were did not have viral load suppression (defined as HIV RNA <1000 copies/mL) and without detectable antiretrovirals (ARVs), were considered indicative of recent infection, with a mean duration of infection of 130 days (95% CI 118–142) in all countries aside from Uganda (153 days, 95% CI 127-178).^19^ We calculated annualized incidence estimates using the World Health Organization (WHO) incidence formula.^20^

We estimated community-level viremia as the weighted proportion of all adults in the sampled enumeration area with a viral load ≥1000 copies/ml, regardless of serostatus and excluding those recently infected to avoid biasing our analysis by including those with the outcome in the exposure variable.^17^

Household wealth quintiles were constructed at the country level using Principal Component Analysis (PCA) based on household assets and infrastructure.^21^

### Statistical analysis

We restricted our analysis to 15-to-59-year-old participants who had been tested for HIV. All analyses were conducted in Stata version 15.1, with Taylor series weighting for variance estimation. All presented percentages and estimates are weighted whereas numbers are crude.

We ran two main analyses, 1) severe food insecurity as the outcome, and 2) recent HIV as the outcome and severe food insecurity as the exposure, using similar methodology. We used logistic regression for model one, retaining in our multivariable model all variables with a p<0.20 in the univariable analysis, then retaining significant variables (p<0.10) in the final model. Goodness of fit of our final model was tested using Hosmer/Lemeshow’s test. We included urbanicity, sex, household wealth quintile, country and age as categorical variables in all models *a priori*.^16 22 23^ For model two, we used Poisson regression due to the rarity of recent HIV as an outcome, to provide the most conservative estimates, and stratified analyses by sex, due to evidence of inequity in impact of severe food insecurity.^12^ We also analyzed mediating behaviors identified in our framework using Poisson regression in a similar fashion to model two, restricted to those reporting ever having sexual activity, aside from the analysis of early sexual debut. We restricted our analysis of intergenerational sex to young women aged 15-24 as these partnerships are particularly risky in this age group.^24 25^ We excluded data from Tanzania in the analysis of forced sex due to the questions on forced sex being asked of a non-representative sample in that country (for details see [appendix p.7]).

We generated maps of the prevalence of HIV infection, viremia, and any food insecurity with SAGA in QGIS version 3.4. We used georeferenced weighted averages at the enumeration area-level, with all cases linked to the centroid of the EA, and kernel density smoothing and interpolation over 200 adult participants for each smoothing circle.

## RESULTS

We enrolled 54,033 households, with 112,955 adults aged 15-59 with HIV test results and data on food insecurity. The majority of heads of households were men, although more women were heads of households in Lesotho (50.9%, n=3621/7502), Eswatini (54.7%, n=2549/4652) and Namibia (51.2%, n=4041/8002, Table 1). Most participants were rural (63.5%, n=73501/54033), with the highest rural proportions in Uganda and Eswatini, and the lowest in Namibia. The largest age group was 15-24 years old, comprising 40.2% (n=42112/112995) of the weighted population. The proportion of participants who had a secondary or greater education was highest in Eswatini (70.6%, n=6477/9553) and Namibia (70.0%, n=9979/16267), and lowest in Tanzania (25.6%, n=6490/28340). Less than half of participants (45.8%, 47357/112995) had been formally employed in the past 12 months. HIV prevalence was highest in women in Eswatini (34.2%, n=1913/5525, Figure 1A), and lowest in men in Tanzania (3.5%, n=521/12297). More HIV-positive men (47.5%, n=1727/4473) than women (37.9%, n=2963/9736) had unsuppressed viral load, which was highest in men in Tanzania (58.7%, n=301/521), and community viremia was highest in Lesotho, although highly heterogeneous across countries (Figure 1B).

**Table 1.** Characteristics of participating households and adults aged 15-59, by country.

| **Characteristic** | **Eswatini**<br>N=4,652<br>% (n) | **Lesotho**<br>N=7,052<br>% (n) | **Tanzania**<br>N=13,328<br>% (n) | **Uganda**<br>N=11,717<br>% (n) | **Namibia**<br>N=8,002<br>% (n) | **Zambia**<br>N=9,282<br>% (n) | **Total**<br>N=54,033<br>% (n) |
| --- | --- | --- | --- | --- | --- | --- | --- |
| Households reporting any food insecurity | 31.3 (1,531) | 31.1 (2,273) | 21.4 (2,818) | 27.8 (3,599) | 22.9 (2,015) | 17.8 (1,628) | 23.5 (13,864) |
| Median Youth Dependency ratio (IQR)<sup>a</sup> | 40 (0-55) | 33 (0-50) | 50 (33-64) | 50 (33-67) | 33 (0-50) | 100 (40-200) | 100 (50-150) |
| Female head of household | 54.7 (2,549) | 50.9 (3,621) | 26.8 (3,417) | 31.0 (3,714) | 51.2 (4,041) | 23.7 (2,196) | 29.2 (19,530) |
| **Receipt of economic support<sup>b</sup>** |  |  |  |  |  |  |  |
| None | 63.9 (2,904) | 79.7 (5,560) | 94.2 (12,521) | 93.7 (10,961) | 71.7 (5,404) | 96.1 (8,923) | 93.1 (46,273) |
| Economic only | 19.1 (914) | 13.6 (981) | 5.2 (736) | 5.0 (593) | 19.7 (1,842) | 3.0 (271) | 5.5 (6,729) |
| Food support | 17.0 (834) | 6.7 (511) | 0.6 (71) | 1.2 (163) | 8.6 (756) | 0.9 (88) | 1.3 (2,423) |
| **Individual level** | **N=9,553**<br>% (n) | **N=11,655**<br>% (n) | **N=28,340**<br>% (n) | **N=28,030**<br>% (n) | **N=16,267**<br>% (n) | **N=19,110**<br>% (n) | **N=112,955**<br>% (n) |
| **Geography - % (n)** |  |  |  |  |  |  |  |
| Urban | 28.0 (2,131) | 48.8 (5,208) | 37.5 (9,348) | 28.8 (7,663) | 58.4 (6,765) | 45.7 (8,339) | 36.5 (39,454) |
| Rural | 72.0 (7,422) | 51.2 (6,447) | 62.5 (18,992) | 71.2 (20,367) | 41.6 (9,502) | 54.3 (10,771) | 63.5 (73,501) |
| **Sex- % (n)** |  |  |  |  |  |  |  |
| Women | 54.4 (5,525) | 49.7 (6,870) | 50.8 (16,043) | 52.5 (16,094) | 51.5 (9,220) | 51.1 (10,981) | 51.4 (64,726) |
| Men | 45.6 (4,028) | 50.3 (4,785) | 49.2 (12,297) | 47.5 (11,945) | 48.5 (7,047) | 48.9 (8,129) | 48.6 (48,229) |
| Median age (IQR) | 28 (21-38) | 30 (22-40) | 28 (20-39) | 27 (20-37) | 29 (21-40) | 27 (20-38) | 28 (20-38) |
| **Age group (years)** |  |  |  |  |  |  |  |
| 15-24 | 37.2 (3,599) | 34.1 (4,037) | 38.5 (10,359) | 43.3 (11,241) | 35.2 (5,557) | 41.1 (7,319) | 40.2 (42,112) |
| 25-34 | 29.0 (2,600) | 29.8 (3,223) | 27.3 (7,704) | 26.9 (7,613) | 28.6 (4,302) | 27.3 (5,130) | 27.3 (30,572) |
| 35-44 | 18.7 (1,698) | 19.0 (2,148) | 18.8 (5,603) | 16.4 (4,879) | 19.4 (3,310) | 18.1 (3,736) | 18.0 (21,374) |
| 45-59 | 15.1 (1,656) | 17.1 (2,247) | 15.4 (4,674) | 13.4 (4,297) | 16.8 (3,098) | 13.5 (2,925) | 14.5 (18,897) |
| **No food in house in past 4 weeks** |  |  |  |  |  |  |  |
| Never | 66.4 (6,089) | 69.0 (7,820) | 79.3 (22,479) | 72.1 (19,441) | 75.6 (11,788) | 83.1 (15,896) | 77.1 (83,513) |
| Rarely (1-2 X) | 18.6 (1,900) | 13.1 (1,569) | 11.3 (3,299) | 15.8 (4,767) | 11.1 (1,944) | 10.2 (1,954) | 12.7 (15,433) |
| Sometimes (3-10X) | 12.3 (1,280) | 13.4 (1,678) | 7.2 (2,010) | 10.7 (3,433) | 10.9 (2,104) | 5.8 (1,089) | 8.4 (11,594) |
| Often (>10x) | 2.8 (284) | 4.5 (588) | 2.2 (552) | 1.4 (389) | 2.4 (431) | 0.9 (171) | 1.8 (2,415) |
| **Educational level** |  |  |  |  |  |  |  |
| None | 3.5 (372) | 5.0 (572) | 12.4 (4,135) | 7.1 (2,451) | 6.7 (1,490) | 5.0 (961) | 9.3 (9,981) |
| Primary | 25.9 (2,722) | 39.7 (4,912) | 62.0 (17,703) | 55.6 (15,882) | 23.3 (4,757) | 41.9 (8,322) | 55.5 (54,298) |
| Secondary or greater | 70.6 (6,447) | 55.3 (6,164) | 25.6 (6,490) | 37.3 (9,538) | 70.0 (9,979) | 53.1 (9,814) | 35.2 (48,432) |
| **Marital status** |  |  |  |  |  |  |  |
| Never married | 55.3 (5,178) | 38.6 (4,267) | 32.1 (7,914) | 33.1 (8,263) | 60.4 (9,228) | 37.2 (6,355) | 34.2 (41,205) |
| Married | 36.8 (3,519) | 48.1 (5,624) | 57.0 (17,086) | 53.6 (15,790) | 32.1 (5,583) | 53.6 (10,648) | 54.5 (58,250) |
| Separated/divorced/widowed | 7.9 (800) | 13.3 (1,743) | 10.9 (3,285) | 13.3 (3,910) | 7.5 (1,310) | 9.3 (1,965) | 11.4 (13,013) |
| **Employed in past 12 months** | 43.3 (3,853) | 39.2 (4,099) | 44.7 (12,155) | 53.2 (14,346) | 45.9 (6,756) | 33.8 (6,148) | 45.8 (47,357) |
| **Recent migrant<sup>c</sup>** | 10.9 (947) | 6.3 (675) | 14.6 (4,005) | 24.5 (6,556) | 29.2 (4,383) | 14.3 (2,683) | 17.9 (19,249) |
| **HIV-positive** |  |  |  |  |  |  |  |
| Women | 34.2 (1,913) | 30.3 (2,161) | 6.5 (1,187) | 7.7 (1,163) | 15.6 (1,623) | 14.6 (1,689) | 9.0 (9,736) |
| Men | 20.5 (875) | 20.9 (1,032) | 3.5 (521) | 4.7 (545) | 9.2 (722) | 9.3 (778) | 5.3 (4,473) |
| **Viral load>1000 copies/ml<sup>d</sup>** |  |  |  |  |  |  |  |
| Women | 24.6 (461) | 29.5 (617) | 42.5 (513) | 37.7 (432) | 18.5 (313) | 39.8 (627) | 37.9 (2,963) |
| Men | 33.8 (274) | 36.4 (364) | 58.7 (301) | 46.1 (247) | 31.1 (220) | 42.5 (321) | 47.5 (1,727) |

Note- some totals may equal greater than 100% due to rounding. Data are survey weighted using Taylor series weights for estimates of variance.
<sup>a</sup> Dependency ratio was calculated as the number of rostered usual residents aged 0-14/(rostered 15 and older)\*100.
<sup>b</sup> Measured over the past 3 months.
<sup>c</sup> Migrant defined as being away from home for at least one month in the past 12 months, except for Namibia, where it was during the past three years.
<sup>d</sup> Among HIV-positive.

**Figure 1:**
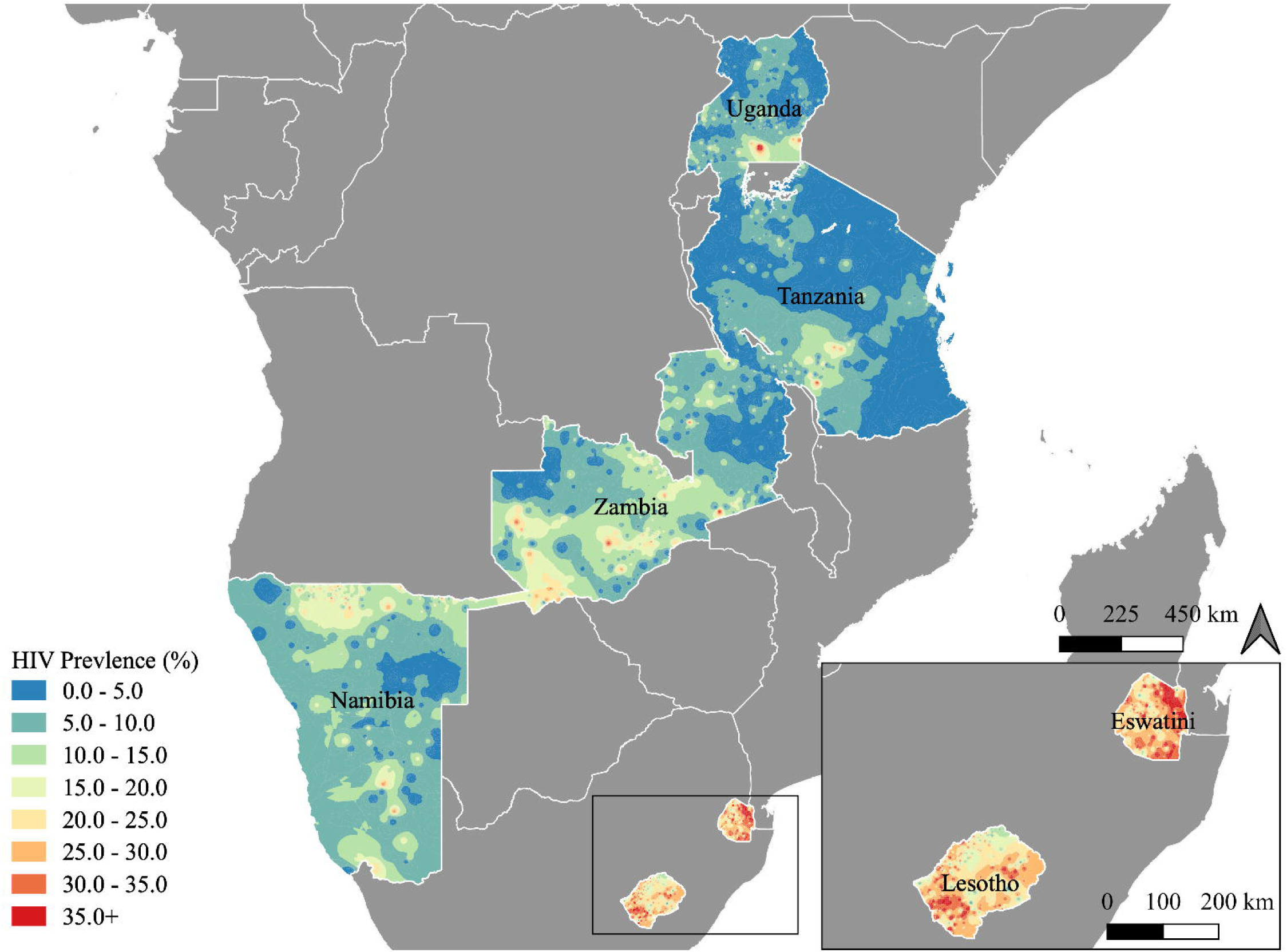
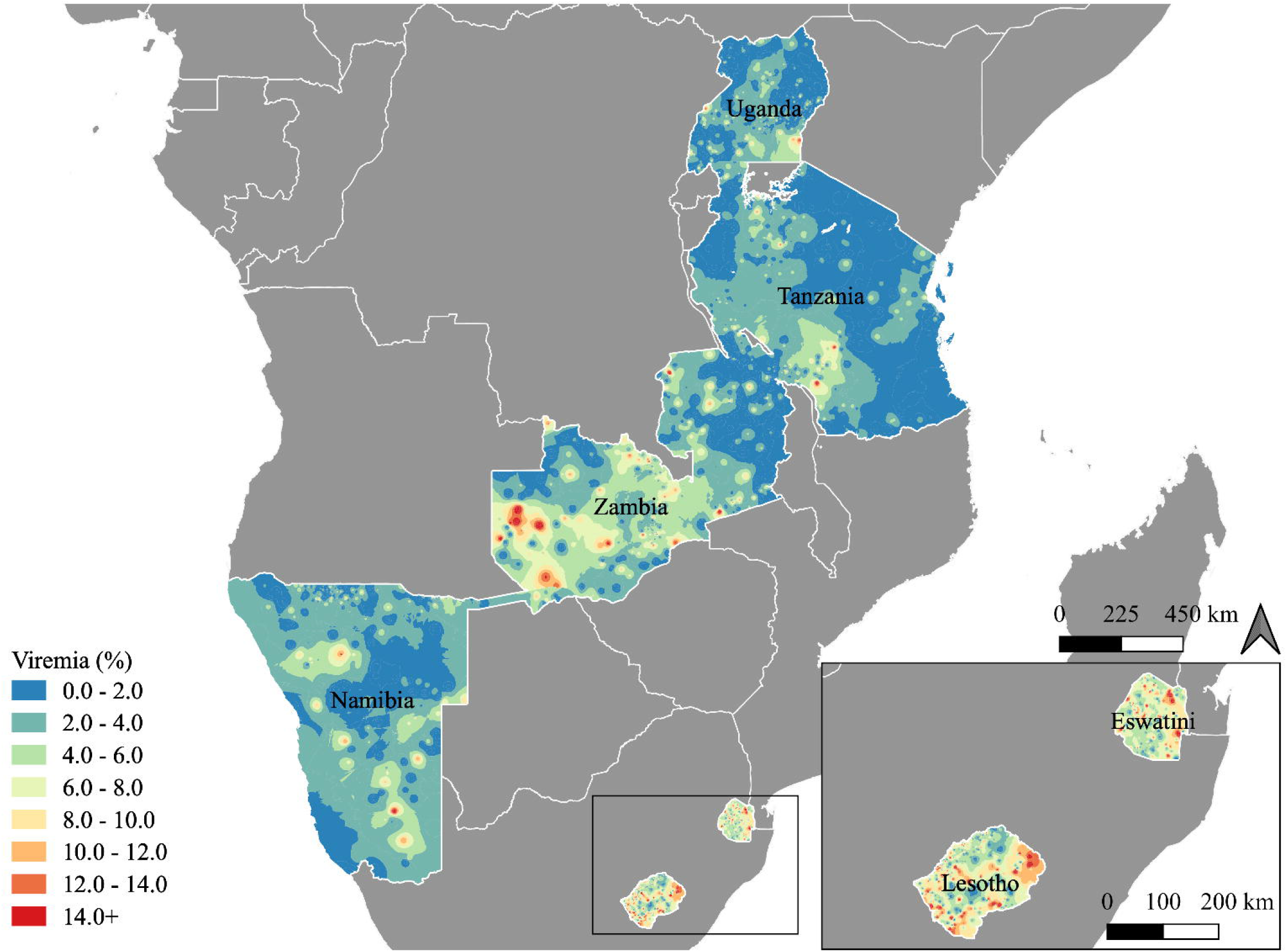
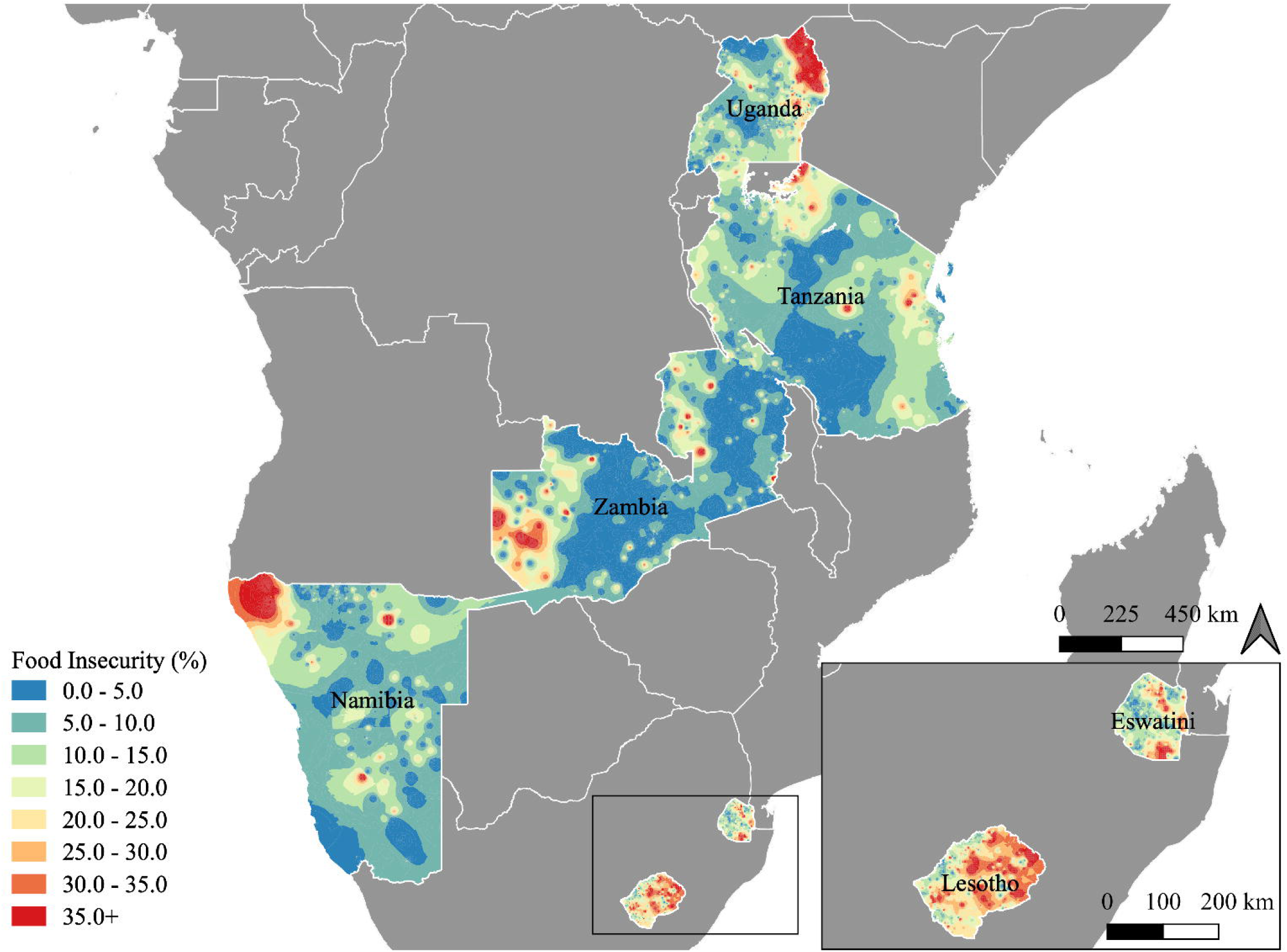
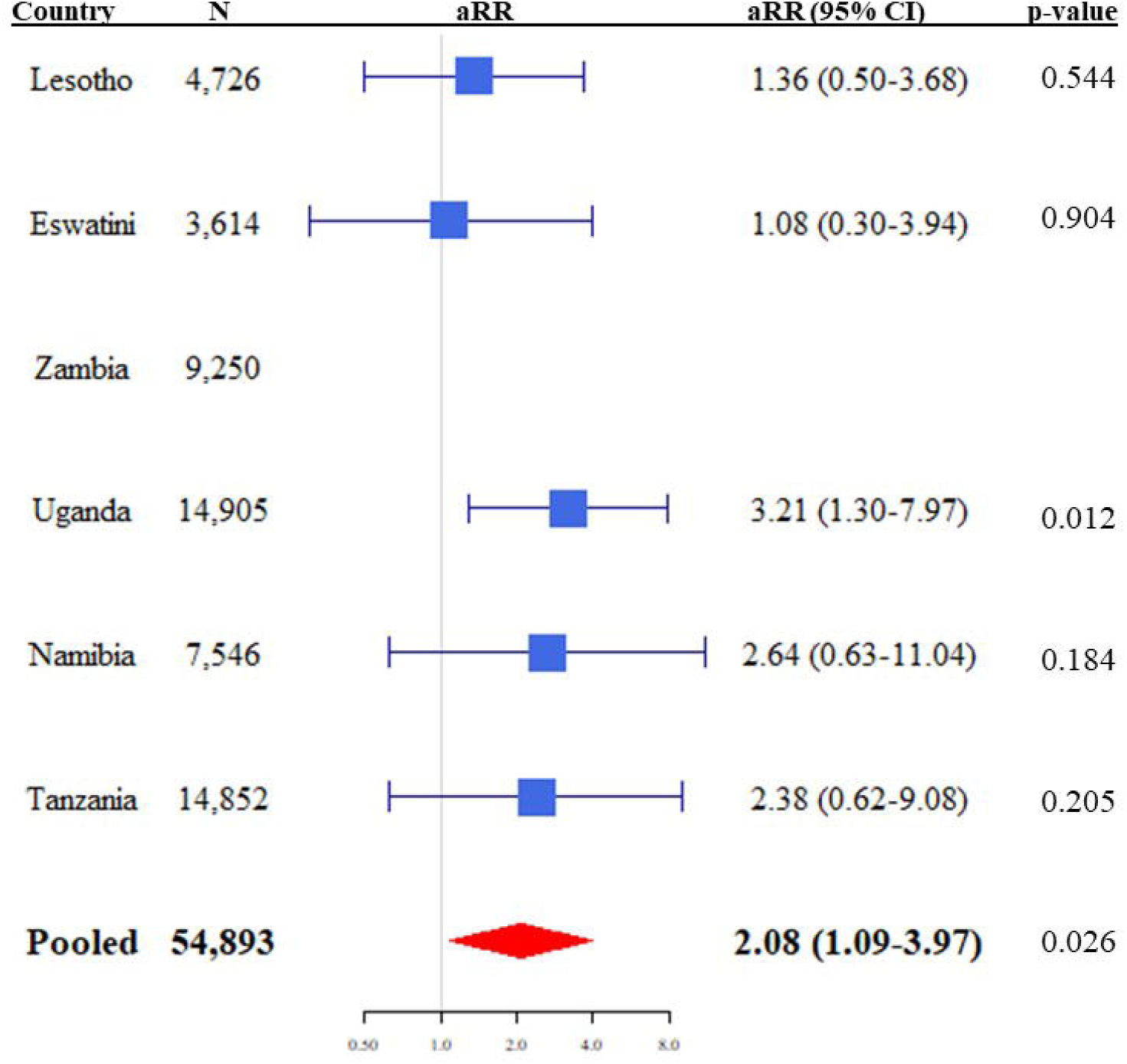
Weighted maps of the prevalence of (A) HIV infection, (B) Community HIV viremia, and (C) Any food insecurity in adults aged 15-59 in six countries in Africa, 2016-2017. Legend: Maps were generated with SAGA in QGIS version 3.4. We used georeferenced weighted averages at the enumeration area-level, with all cases linked to the centroid of the EA, and kernel density smoothing and interpolation over 200 adult participants for each smoothing circle. **(B)** community HIV viremia (%) was defined as a viral load >1000 copies/ml, in the total sampled population regardless of HIV serostatus; (C) any food insecurity defined as no food in the house at least once in the past four weeks.

### Correlates of severe food insecurity

Across all countries, 23.5% (n=13864/54033) of households reported having experienced any lack of food in the past 4 weeks, with 10.2% (n=14009/112955) of participants reporting severe food insecurity. All countries had regions with high burdens of food insecurity, but the distribution was highly heterogeneous, with frequency of any food insecurity ranging from 0-80% of an enumeration area’s population (Figure 1C). The highest prevalence of severe food insecurity was seen in Lesotho (17.9%, 2266/11655, Table 2). Adjusted results from the multivariable analysis were similar to univariable results: male-headed households were less likely to suffer from severe food insecurity (aOR 0.71, 95% CI 0.63-0.80), as were wealthier households (aOR 0.68, 95% CI 0.64-0.71 for each quintile increase in wealth). At the individual level, secondary or greater education (aOR 0.75, 95% CI 0.68-0.82), formal employment in the past year (aOR 0.90, 95% CI 0.84-0.97), and being married (aOR 0.81, 95% CI 0.71-0.91 compared to never married) were all protective against severe food insecurity. Living in a household with many young dependents (aOR 1.12, 95% CI 1.08-1.17 per quartile increase), being aged 35-44 or 45-59 compared to 15-24, being separated/divorced or widowed (aOR 1.17, 95% 1.02-1.33), having recently migrated (aOR 1.14, 95% CI 1.05-1.24), and being HIV-positive were all associated with severe food insecurity (aOR 1.23, 95% CI 1.10-1.38). Excluding those who were recently HIV-infected did not change the latter association. After adjustment, sex, receipt of social support, and HIV-status of the head of household were no longer significant, and rural residence became protective.

**Table 2.**
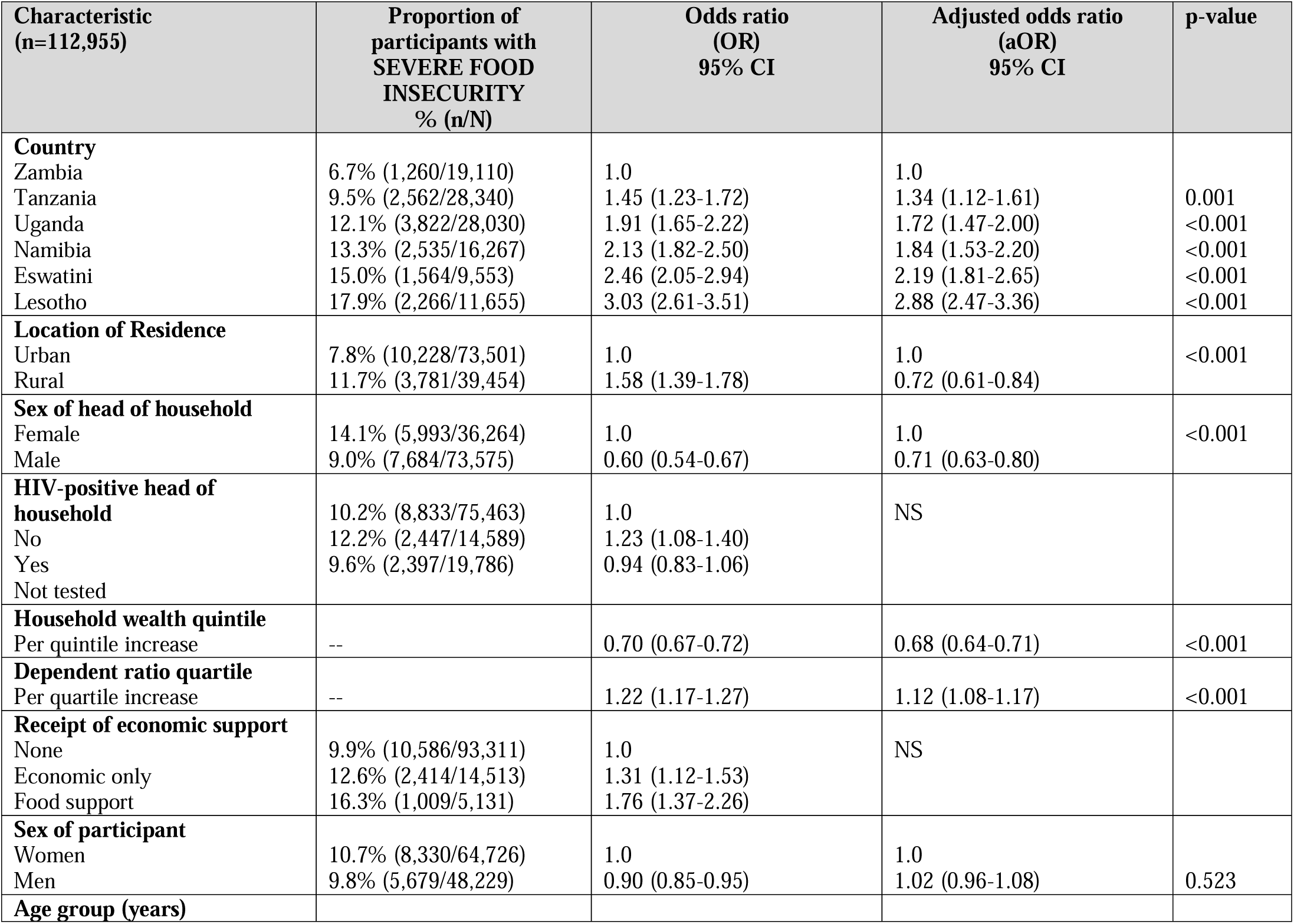
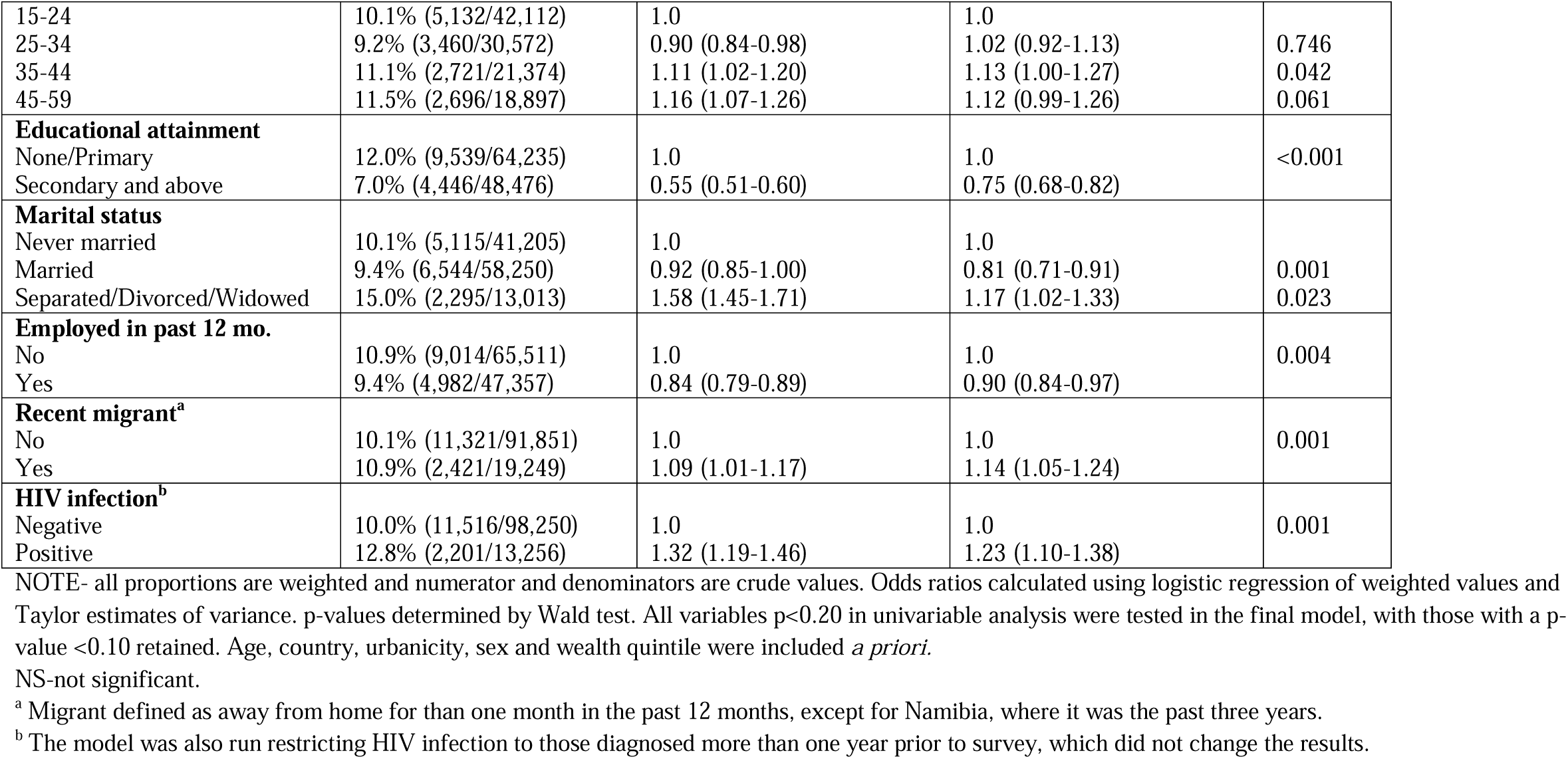
Correlates of severe food insecurity among adults aged 15-59.

| **Characteristic**<br>**(n=112,955)** | **Proportion of**<br>**participants with**<br>**SEVERE FOOD**<br>**INSECURITY**<br>**% (n/N)** | **Odds ratio**<br>**(OR)**<br>**95% CI** | **Adjusted odds ratio**<br>**(aOR)**<br>**95% CI** | **p-value** |
| --- | --- | --- | --- | --- |
| **Country** |  |  |  |  |
| Zambia | 6.7% (1,260/19,110) | 1.0 | 1.0 |  |
| Tanzania | 9.5% (2,562/28,340) | 1.45 (1.23-1.72) | 1.34 (1.12-1.61) | 0.001 |
| Uganda | 12.1% (3,822/28,030) | 1.91 (1.65-2.22) | 1.72 (1.47-2.00) | <0.001 |
| Namibia | 13.3% (2,535/16,267) | 2.13 (1.82-2.50) | 1.84 (1.53-2.20) | <0.001 |
| Eswatini | 15.0% (1,564/9,553) | 2.46 (2.05-2.94) | 2.19 (1.81-2.65) | <0.001 |
| Lesotho | 17.9% (2,266/11,655) | 3.03 (2.61-3.51) | 2.88 (2.47-3.36) | <0.001 |
| **Location of Residence** |  |  |  |  |
| Urban | 7.8% (10,228/73,501) | 1.0 | 1.0 | <0.001 |
| Rural | 11.7% (3,781/39,454) | 1.58 (1.39-1.78) | 0.72 (0.61-0.84) |  |
| **Sex of head of household** |  |  |  |  |
| Female | 14.1% (5,993/36,264) | 1.0 | 1.0 | <0.001 |
| Male | 9.0% (7,684/73,575) | 0.60 (0.54-0.67) | 0.71 (0.63-0.80) |  |
| **HIV-positive head of household** |  |  |  |  |
| No | 10.2% (8,833/75,463) | 1.0 | NS |  |
| Yes | 12.2% (2,447/14,589) | 1.23 (1.08-1.40) |  |  |
| Not tested | 9.6% (2,397/19,786) | 0.94 (0.83-1.06) |  |  |
| **Household wealth quintile** |  |  |  |  |
| Per quintile increase | -- | 0.70 (0.67-0.72) | 0.68 (0.64-0.71) | <0.001 |
| **Dependent ratio quartile** |  |  |  |  |
| Per quartile increase | -- | 1.22 (1.17-1.27) | 1.12 (1.08-1.17) | <0.001 |
| **Receipt of economic support** |  |  |  |  |
| None | 9.9% (10,586/93,311) | 1.0 | NS |  |
| Economic only | 12.6% (2,414/14,513) | 1.31 (1.12-1.53) |  |  |
| Food support | 16.3% (1,009/5,131) | 1.76 (1.37-2.26) |  |  |
| **Sex of participant** |  |  |  |  |
| Women | 10.7% (8,330/64,726) | 1.0 | 1.0 |  |
| Men | 9.8% (5,679/48,229) | 0.90 (0.85-0.95) | 1.02 (0.96-1.08) | 0.523 |
| **Age group (years)** |  |  |  |  |
| 15-24 | 10.1% (5,132/42,112) | 1.0 | 1.0 |  |
| 25-34 | 9.2% (3,460/30,572) | 0.90 (0.84-0.98) | 1.02 (0.92-1.13) | 0.746 |
| 35-44 | 11.1% (2,721/21,374) | 1.11 (1.02-1.20) | 1.13 (1.00-1.27) | 0.042 |
| 45-59 | 11.5% (2,696/18,897) | 1.16 (1.07-1.26) | 1.12 (0.99-1.26) | 0.061 |
| **Educational attainment** |  |  |  |  |
| None/Primary | 12.0% (9,539/64,235) | 1.0 | 1.0 | <0.001 |
| Secondary and above | 7.0% (4,446/48,476) | 0.55 (0.51-0.60) | 0.75 (0.68-0.82) |  |
| **Marital status** |  |  |  |  |
| Never married | 10.1% (5,115/41,205) | 1.0 | 1.0 |  |
| Married | 9.4% (6,544/58,250) | 0.92 (0.85-1.00) | 0.81 (0.71-0.91) | 0.001 |
| Separated/Divorced/Widowed | 15.0% (2,295/13,013) | 1.58 (1.45-1.71) | 1.17 (1.02-1.33) | 0.023 |
| **Employed in past 12 mo.** |  |  |  |  |
| No | 10.9% (9,014/65,511) | 1.0 | 1.0 | 0.004 |
| Yes | 9.4% (4,982/47,357) | 0.84 (0.79-0.89) | 0.90 (0.84-0.97) |  |
| **Recent migrant<sup>a</sup>** |  |  |  |  |
| No | 10.1% (11,321/91,851) | 1.0 | 1.0 | 0.001 |
| Yes | 10.9% (2,421/19,249) | 1.09 (1.01-1.17) | 1.14 (1.05-1.24) |  |
| **HIV infection<sup>b</sup>** |  |  |  |  |
| Negative | 10.0% (11,516/98,250) | 1.0 | 1.0 | 0.001 |
| Positive | 12.8% (2,201/13,256) | 1.32 (1.19-1.46) | 1.23 (1.10-1.38) |  |

NOTE- all proportions are weighted and numerator and denominators are crude values. Odds ratios calculated using logistic regression of weighted values and Taylor estimates of variance. p-values determined by Wald test. All variables $p < 0.20$ in univariable analysis were tested in the final model, with those with a p-value $< 0.10$ retained. Age, country, urbanicity, sex and wealth quintile were included *a priori*.
NS-not significant.
<sup>a</sup> Migrant defined as away from home for than one month in the past 12 months, except for Namibia, where it was the past three years.
<sup>b</sup> The model was also run restricting HIV infection to those diagnosed more than one year prior to survey, which did not change the results.

### Association between severe food insecurity, food and economic support and HIV-related outcomes

Of the 14,208 HIV-positive participants, 1.9% (n=200) were classified as having recent HIV infection, of which 140 were women and 60 were men. Incidence was highest in women aged 15-49 in Eswatini (1.73%, 95% CI 0.96-2.50, Supplementary Figure 6), and lowest in men aged 15-24 in Tanzania (0%, 95% CI 0-0.23). Among those without chronic HIV infection, there were 27 recent cases in 6,699 severely food insecure women, and 113 cases in the other 48,431 women; there were 13 recent cases in 4,974 severely food insecure men, and 47 in the other 38,842 men. In univariable analysis of predictors of recent HIV infection, the relative risk of new infection was highest in women aged 25-34 and in men aged 35-44, and 45-59, compared to participants aged 15-24 (Table 3). Results from our multivariable model demonstrated that severe food insecurity was associated with a two-fold increase in risk of recent infection in women (aRR 2.08, 95% CI 1.09-3.97), with the effect relatively homogeneous across countries, although a lower risk was seen in Lesotho and Eswatini (Figure 2). There was no significant risk noted in men. Both sexes were at higher risk of HIV acquisition if previously married, compared to never married, but currently married men were also at significantly higher risk of recent HIV infection (aRR 8.96, 95% CI 1.77-45.35). Receipt of food support was associated with a pronounced lower risk of recent HIV in women (aRR 0.36, 95% CI 0.14-0.88), whereas receipt of other types of support was not, and neither were protective in men. The use of a scale of food insecurity incorporating the three questions on food availability and access, did not produce substantially different results than our measure [(Supplementary Table 1)], although fewer people were classified as severely food insufficient. This is further discussed in the appendix [(p 9)].

**Table 3.** Multivariable analysis of relative risk of recent HIV infection in those with severe food insecurity in participants aged 15-59, by sex.

| Characteristic | Women<br>(n=54,834) |  |  |  | Men<br>(n=43,827) |  |  |  |
| --- | --- | --- | --- | --- | --- | --- | --- | --- |
|  | RR (95% CI) | P-Value | aRR<br>(95% CI) | P-value | RR<br>(95% CI) | P-Value | aRR<br>(95% CI) | P-value |
| **Severe food insecurity** | **2.11 (1.11-4.03)** | **0.023** | **2.08 (1.09-3.97)** | **0.026** | 1.85 (0.82-4.20) | 0.140 | 1.77 (0.84-3.74) | 0.134 |
| **Age group (years)** |  |  |  |  |  |  |  |  |
| 15-24 | 1.0 |  | 1.0 |  | 1.0 |  | 1.0 |  |
| 25-34 | 1.72 (0.97-3.05) | 0.064 | 1.20 (0.61-2.35) | 0.594 | 2.62 (0.93-7.36) | 0.067 | 0.81 (0.22-3.05) | 0.760 |
| 35-44 | 1.14 (0.59-2.21) | 0.698 | 0.77 (0.35-1.67) | 0.500 | **5.40 (1.92-15.21)** | **0.001** | 1.45 (0.35-5.99) | 0.605 |
| 45-59 | 0.61 (0.21-1.76) | 0.355 | 0.34 (0.11-1.12) | 0.077 | **4.75 (1.49-15.13)** | **0.009** | 1.27 (0.30-5.54) | 0.747 |
| **Country** |  |  |  |  |  |  |  |  |
| Zambia | 1.0 |  | 1.0 |  | 1.0 |  | 1.0 |  |
| Tanzania | 0.38 (0.22-0.66) | 0.001 | **0.46 (0.26-0.82)** | **0.009** | 0.60 (0.24-1.50) | 0.272 | 0.58 (0.22-1.52) | 0.263 |
| Uganda | 0.59 (0.36-0.97) | 0.036 | 0.68 (0.41-1.14) | 0.140 | 1.45 (0.58-3.61) | 0.421 | 1.31 (0.50-3.42) | 0.583 |
| Namibia | 0.67 (0.34-1.30) | 0.233 | 0.89 (0.43-1.83) | 0.749 | 0.47 (0.13-1.67) | 0.243 | 0.72 (0.19-2.73) | 0.631 |
| Eswatini | 1.60 (0.91-2.81) | 0.102 | 1.64 (0.89-3.02) | 0.115 | **3.07 (1.18-8.00)** | **0.022** | **2.84 (1.04-7.79)** | **0.042** |
| Lesotho | 1.32 (0.75-2.30) | 0.331 | 0.86 (0.47-1.56) | 0.614 | **3.46 (1.42-8.47)** | **0.007** | 2.60 (0.99-6.85) | 0.053 |
| **Location of Residence** |  |  |  |  |  |  |  |  |
| Urban | 1.0 |  |  |  | **1.0** |  | 1.0 |  |
| Rural | 0.72 (0.46-1.13) | 0.154 |  |  | **2.44 (1.14-5.25)** | **0.022** | **2.47 (1.14-5.35)** | **0.022** |
| **Wealth Quintile**<br>(per quintile increase) | 0.99 (0.84-1.16) | 0.886 |  |  | **0.81 (0.66-0.98)** | **0.033** |  |  |
| **Community viremia**<br>(per 1% increase) | **1.12 (1.09-1.16)** | **<0.001** | **1.10 (1.05-1.15)** | **<0.001** | **1.10 (1.05-1.16)** | **<0.001** | **1.10 (1.02-1.18)** | **0.015** |
| **Receipt of economic support** |  |  |  |  |  |  |  |  |
| None | 1.0 |  | 1.0 |  | 1.0 |  |  |  |
| Economic only | 1.14 (0.58-2.21) | 0.709 | 1.06 (0.54-2.07) | 0.864 | 1.48 (0.55-3.99) | 0.436 |  |  |
| Food support | 0.51 (0.20-1.32) | 0.162 | **0.36 (0.14-0.88)** | **0.025** | 4.10 (0.74-22.76) | 0.106 |  |  |
| **Migration** |  |  |  |  |  |  |  |  |
| None | 1.0 |  |  |  | 1.0 |  |  |  |
| Away for >1 month | 1.11 (0.57-2.17) | 0.751 |  |  | 0.79 (0.32-1.92) | 0.597 |  |  |
| **Employment status** |  |  |  |  |  |  |  |  |
| No formal employment | 1.0 |  |  |  | 1.0 |  | 1.0 |  |
| Worked in past year | 1.48 (0.93-2.36) | 0.097 |  |  | **2.33 (1.11-4.91)** | **0.026** | 1.95 (0.95-4.02) | 0.070 |
| **Marital status** |  |  |  |  |  |  |  |  |
| Never married | 1.0 |  | 1.0 |  | 1.0 |  | 1.0 |  |
| Married | 1.51 (0.86-2.65) | 0.148 | 1.86 (0.94-3.68) | 0.074 | **10.83 (3.86-30.38)** | **<0.001** | **8.96 (1.77-45.35)** | **0.008** |
| Separated/Divorced/Widowed | 3.10 (1.69-5.71) | <0.001 | **4.25 (1.89-9.57)** | **0.001** | **11.19 (2.21-56.73)** | **0.004** | **8.23 (1.15-59.02)** | **0.036** |
| **Male circumcised** |  |  |  |  | 0.60 (0.30-1.19) | 0.144 | NS | NS |

RR: Relative risk, aRR: adjusted relative risk, CI: confidence interval.
Note- RR determined by Poisson regression using weighted values and Taylor estimates of variance. All variables $p < 0.20$ in univariable analysis were tested in the final model, with those with a $p$ -value $< 0.10$ retained. Age group and country were included *a priori*. Results indicated in bold are significant at $p < 0.05$ .

The frequency of different potential mediating behaviors is described in Supplementary Table 2. Women in Uganda reported the highest frequency of transactional sex (19.5%, n=2,353/11824), early sexual debut (12.1%, n=2,009/15,813), and forced sex (16.2%, n=384/2898), whereas women in Tanzania reported more high-risk sex (45.8%, n=5,038/11,246) and intergenerational sex in young women (18.2%, n=616/3,556). There was a statistically significant association between severe food insecurity and transactional sex (aRR 1.28, 95% CI 1.17-1.41, Table 4). Women with severe food insecurity also reported more frequent early sexual debut (aRR 1.18, 95% CI 1.06-1.31), more forced sex (aRR 1.36, 95% CI 1.11-1.66), and more high-risk sex (aRR 1.08, 95% CI 1.02-1.14). Economic (aRR 0.89, 95% CI 0.84-0.95) and food support (aRR 0.81, 95% CI 0.69-0.97) were both associated with significantly lower risks of high-risk sex. Severe food insecurity was also associated with an elevated risk (aRR 1.23, 95% 1.03-1.46) of intergenerational sex, reported by 16.5% of young women. None of the behaviors were associated with urbanicity after adjusting for other demographic factors. There was heterogeneity between countries for the increased risk of forced and intergenerational sex in food insecure women (Supplementary Figure 7).

**Table 4.**
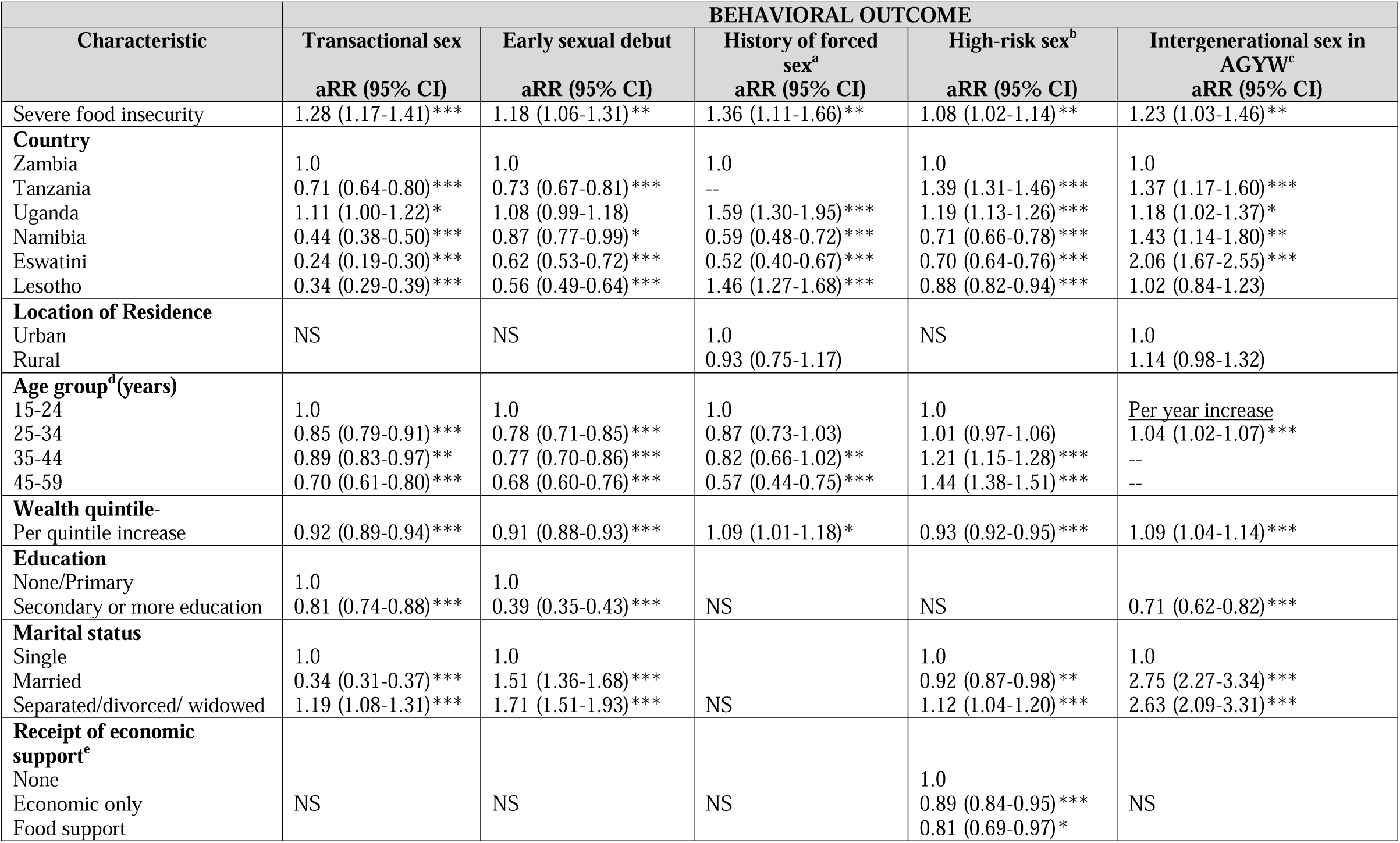
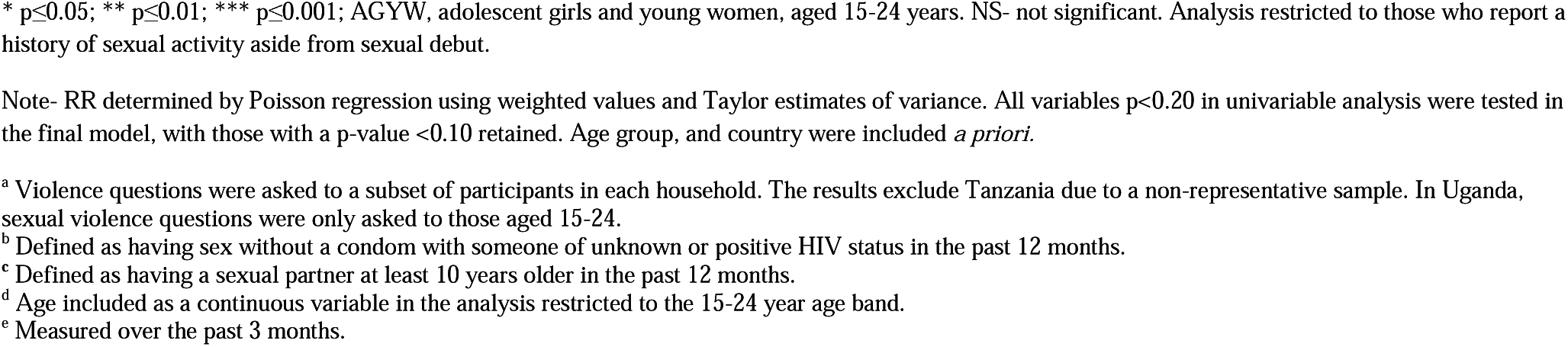
Multivariable analysis of the relative risk of several high-risk sexual behaviors among women aged 15-59 with severe food insecurity.

| Characteristic | BEHAVIORAL OUTCOME |  |  |  |  |
| --- | --- | --- | --- | --- | --- |
|  | Transactional sex<br>aRR (95% CI) | Early sexual debut<br>aRR (95% CI) | History of forced sex<sup>a</sup><br>aRR (95% CI) | High-risk sex<sup>b</sup><br>aRR (95% CI) | Intergenerational sex in AGYW<sup>c</sup><br>aRR (95% CI) |
| Severe food insecurity | 1.28 (1.17-1.41)*** | 1.18 (1.06-1.31)** | 1.36 (1.11-1.66)** | 1.08 (1.02-1.14)** | 1.23 (1.03-1.46)** |
| **Country** |  |  |  |  |  |
| Zambia | 1.0 | 1.0 | 1.0 | 1.0 | 1.0 |
| Tanzania | 0.71 (0.64-0.80)*** | 0.73 (0.67-0.81)*** | -- | 1.39 (1.31-1.46)*** | 1.37 (1.17-1.60)*** |
| Uganda | 1.11 (1.00-1.22)* | 1.08 (0.99-1.18) | 1.59 (1.30-1.95)*** | 1.19 (1.13-1.26)*** | 1.18 (1.02-1.37)* |
| Namibia | 0.44 (0.38-0.50)*** | 0.87 (0.77-0.99)* | 0.59 (0.48-0.72)*** | 0.71 (0.66-0.78)*** | 1.43 (1.14-1.80)** |
| Eswatini | 0.24 (0.19-0.30)*** | 0.62 (0.53-0.72)*** | 0.52 (0.40-0.67)*** | 0.70 (0.64-0.76)*** | 2.06 (1.67-2.55)*** |
| Lesotho | 0.34 (0.29-0.39)*** | 0.56 (0.49-0.64)*** | 1.46 (1.27-1.68)*** | 0.88 (0.82-0.94)*** | 1.02 (0.84-1.23) |
| **Location of Residence** |  |  |  |  |  |
| Urban | NS | NS | 1.0 | NS | 1.0 |
| Rural |  |  | 0.93 (0.75-1.17) |  | 1.14 (0.98-1.32) |
| **Age group<sup>d</sup>(years)** |  |  |  |  |  |
| 15-24 | 1.0 | 1.0 | 1.0 | 1.0 | <u>Per year increase</u> |
| 25-34 | 0.85 (0.79-0.91)*** | 0.78 (0.71-0.85)*** | 0.87 (0.73-1.03) | 1.01 (0.97-1.06) | 1.04 (1.02-1.07)*** |
| 35-44 | 0.89 (0.83-0.97)** | 0.77 (0.70-0.86)*** | 0.82 (0.66-1.02)** | 1.21 (1.15-1.28)*** | -- |
| 45-59 | 0.70 (0.61-0.80)*** | 0.68 (0.60-0.76)*** | 0.57 (0.44-0.75)*** | 1.44 (1.38-1.51)*** | -- |
| **Wealth quintile-**<br>**Per quintile increase** | 0.92 (0.89-0.94)*** | 0.91 (0.88-0.93)*** | 1.09 (1.01-1.18)* | 0.93 (0.92-0.95)*** | 1.09 (1.04-1.14)*** |
| **Education** |  |  |  |  |  |
| None/Primary | 1.0 | 1.0 |  |  |  |
| Secondary or more education | 0.81 (0.74-0.88)*** | 0.39 (0.35-0.43)*** | NS | NS | 0.71 (0.62-0.82)*** |
| **Marital status** |  |  |  |  |  |
| Single | 1.0 | 1.0 |  | 1.0 | 1.0 |
| Married | 0.34 (0.31-0.37)*** | 1.51 (1.36-1.68)*** |  | 0.92 (0.87-0.98)** | 2.75 (2.27-3.34)*** |
| Separated/divorced/ widowed | 1.19 (1.08-1.31)*** | 1.71 (1.51-1.93)*** | NS | 1.12 (1.04-1.20)*** | 2.63 (2.09-3.31)*** |
| **Receipt of economic support<sup>e</sup>** |  |  |  |  |  |
| None |  |  |  | 1.0 |  |
| Economic only | NS | NS | NS | 0.89 (0.84-0.95)*** | NS |
| Food support |  |  |  | 0.81 (0.69-0.97)* |  |

\* $p \leq 0.05$ ; \*\* $p \leq 0.01$ ; \*\*\* $p \leq 0.001$ ; AGYW, adolescent girls and young women, aged 15-24 years. NS- not significant. Analysis restricted to those who report a history of sexual activity aside from sexual debut.
Note- RR determined by Poisson regression using weighted values and Taylor estimates of variance. All variables $p < 0.20$ in univariable analysis were tested in the final model, with those with a p-value $< 0.10$ retained. Age group, and country were included *a priori*.
<sup>a</sup> Violence questions were asked to a subset of participants in each household. The results exclude Tanzania due to a non-representative sample. In Uganda, sexual violence questions were only asked to those aged 15-24.
<sup>b</sup> Defined as having sex without a condom with someone of unknown or positive HIV status in the past 12 months.
<sup>c</sup> Defined as having a sexual partner at least 10 years older in the past 12 months.
<sup>d</sup> Age included as a continuous variable in the analysis restricted to the 15-24 year age band.
<sup>e</sup> Measured over the past 3 months.

## DISCUSSION

To our knowledge, this is the first study to directly link acute food insecurity with HIV incidence in women in sub-Saharan Africa, supporting prior studies which have shown associations between hunger, sexual risk-taking, and prevalent HIV infection.^11 12^,^22^ The robustness and representativeness of the PHIA data, spanning multiple countries and contexts, including highly variable community HIV burden, is particularly valuable for substantiating the likely pathways for this association.^12 26^ Our data also suggest that some of the communities with the highest levels of any food insecurity, such as areas in Lesotho, tend to have the highest HIV prevalence; these findings could therefore have serious implications for efforts to achieve or maintain epidemic control.

There was substantial variation in the spatial distribution of households reporting any food insecurity, both across countries and sub-nationally. However, we were able to find consistent correlates with insecurity: poorer households headed by women, or with many children, had much higher odds of severe food shortages. These findings have been shown in multiple other studies, attributed to the fact that women often have little control over resources such as land and employment, leading to a disproportionate susceptibility to poverty and income shocks.^27^ It is noteworthy that female sex is not significant in our adjusted model, and that marriage is protective, suggesting that women in male-headed-households are protected by their husband’s income-earning potential. The bidirectionality of the relationship between food insecurity and HIV infection is also seen here, where infection was strongly associated with severe food insecurity even when we restricted our analysis to those infected for more than one year.^16^

The two-fold increase in risk of recent HIV infection seen in women who reported severe food insecurity reinforces other studies which have shown increases in risk behavior and higher HIV prevalence in food insecure women,^14 28 29^ but allows us to better understand the direction of the association. This increased acquisition may be attributed to the constellation of risk factors impacting these women, including more transactional sex. The risk of transactional sex declined with age, and with wealth and education. These findings build on previous studies which found that food insecurity and poverty is commonly associated with sex in exchange for goods.^12 22 26 27^ Young food-insecure women were also more likely to report significantly older partners, possibly because they confer some financial benefit. These older partners are often more infectious than same-aged partners as a result of higher rates of viremia due to having recently acquired HIV, being undiagnosed, or not taking ART.^24 25 30^ It is also noteworthy that women of all ages compounded their risk by not using condoms with men who might be HIV positive, a risk factor implicated as a key driver of HIV acquisition.^31^ Finally, the fact that food-insecure women reported both more forced sex over their lifetimes, as well as more frequent early sexual debut suggests that some of the risk behaviors are a result of compounded vulnerabilities, and that some experiences might precede or predispose women to food insecurity. These findings also support that food security has a significant gender dimension, where women are both more at risk of severe hunger, and suffer more consequences due to limited coping strategies, which includes different forms of sexual activity in exchange for material support.^14^ Finally, the inter-country heterogeneity of certain risk behaviors suggests that there might be multiple different pathways between food insecurity and HIV acquisition, and these are likely to be highly contextual.

While most forms of social support were not associated with a protective effect, receipt of food support was associated with a 64% lower risk of recent HIV infection in women. This suggests that hunger alleviation interrupts the cycle of vulnerability, possibly because food support generally goes directly to women who are responsible for intra-household consumption needs, and is unlikely to be used by men for other purchases.^32^ Our results suggest that food support addresses women’s immediate food shortage, alleviating the pressure to engage in forms of high-risk behaviors to obtain food. Further analysis of our data, disaggregated by sex, age and risk group, and epidemiological context, is currently being conducted to understand how different forms of social support impact behaviors and HIV risk; this should enable the comparison of our data with other studies which have shown benefits of social or financial support, both in terms of short-term assistance and longer-term coping strategies.^33 34^ This research supports the need to address structural constraints underlying poverty, as well as behavioral change and gender equity, and underscores the importance of including women as active agents who can assist in understanding how best to use social assistance.

Limitations of this study include the single-point estimate of HIV infection and associated behaviors, where the cross-sectional nature of the data means that the direction of effect is difficult to determine with certainty. The LAg avidity assay has limitation in estimation of HIV incidence and the algorithm excludes anyone who might have started antiretroviral drugs within the first six months of infection.^20^ We also had relatively small numbers of people classified as recently infected across the surveys, particularly men, which prohibited an in-depth analysis of factors driving HIV infection in this group. However, in women, our findings are supported by our theoretical framework, suggesting that the findings are robust. The attenuation of any effect seen in the multivariable model of recent HIV infection in men also suggests that the patterns of risk for this sex are more context dependent, and therefore a pooled analysis across several different countries is less appropriate. Furthermore, as hazardous drinking data were not collected in all PHIA countries, it prohibited its inclusion in our models, which might have obscured its importance as a driver of both food shortages as well as HIV acquisition.^35^ Further research incorporating more community level variables, as well as other factors which might be more predictive of infection in males, are critical to the identification of high-risk men.

## Conclusions

In conclusion, in this time of global economic disruption and stark increases in food insecurity, it is critical to support the economic empowerment of women, but also the more immediate targeting of food support to the communities most vulnerable to the devastating effects of climate change. Understanding that food insecurity has both short and long-term consequences, including HIV transmission, should spur further investments in preparedness, including in crop and community resilience and environmental justice. The international recognition that food support prevents conflict is heartening, but global donors must also consider other consequences of hunger, including the risks to HIV epidemic control in communities with ongoing high incidence compounded by poverty and food shortages.

## Data Availability

All data used in this manuscript are publicly available at https://phia-data.icap.columbia.edu/.

## Acknowledgments & Funding

This project has been supported by the President’s Emergency Plan for AIDS Relief (PEPFAR) through the Centers for Disease Control and Prevention (CDC) under the terms of cooperative agreement #U2GGH001226. The findings and conclusions in this manuscript are those of the authors and do not necessarily represent the official position of the funding agencies. Co-authors from the CDC participated in study design, data collection, analysis, and interpretation, and in the writing of the report. The corresponding author had final responsibility for the decision to submit for publication.

The authors would also like to acknowledge all PHIA participants and survey staff, and the Ministries of Health of Zambia, Lesotho, Eswatini, Namibia, Tanzania and Uganda, and colleagues from the PHIA research group for their contributions to this study.

## Competing interests

The authors declare that they have no competing interests.

## Authors’ contributions

AL, EG, AS, NM, JJ, CG, HP, AH, WK, KJ, SF, and JW designed the study. AL, EG, AS, NM, JJ, CG, SB, KS, AH, DB, JM, WK, NP, LM, JW, and SF designed the data collection tools. AL and EG did the statistical analysis. AL took primary responsibility for writing the manuscript. All co-authors contributed to data analysis and interpretation, and to the writing and review of the manuscript.

## Supplementary File

Additional file 1: Food_Insecurity_Appendix_09_2021.docx.

This appendix contains additional details on the analytic framework, study design, variable construction, the climate context at the time of data collection, and more information on the measurement of food insecurity.

## Notes

### Competing Interest Statement

The authors have declared no competing interest.

### Clinical Protocols

https://phia.icap.columbia.edu/

### Funding Statement

This project has been supported by the President Emergency Plan for AIDS Relief (PEPFAR) through the Centers for Disease Control and Prevention (CDC) under the terms of cooperative agreement #U2GGH001226.

### Author Declarations

The PHIA protocol and data collection tools were approved by national ethics committees for each country, and the institutional review boards at Columbia University Irving Medical Center, the US Centers for Disease Control and Prevention (CDC) and the University of California, San Francisco in the case of Namibia. Due to the inclusion of six countries and the multiple ethical boards involved, we are providing the protocol numbers for the Columbia University Irving Medical Center, which approved all protocols (AAAQ0753, AAAQ7860, AAAQ8408, AAAQ8537, AAAR2051, AAAQ889).

