## Supplemental file for "Food insecurity and the risk of HIV acquisition: Findings from population-based surveys in six sub-Saharan African countries (2016-2017)"

**Supplementary Material**

### Theoretical framework for relationship between climate change and HIV

#### Supplementary Figure 1. Theoretical framework

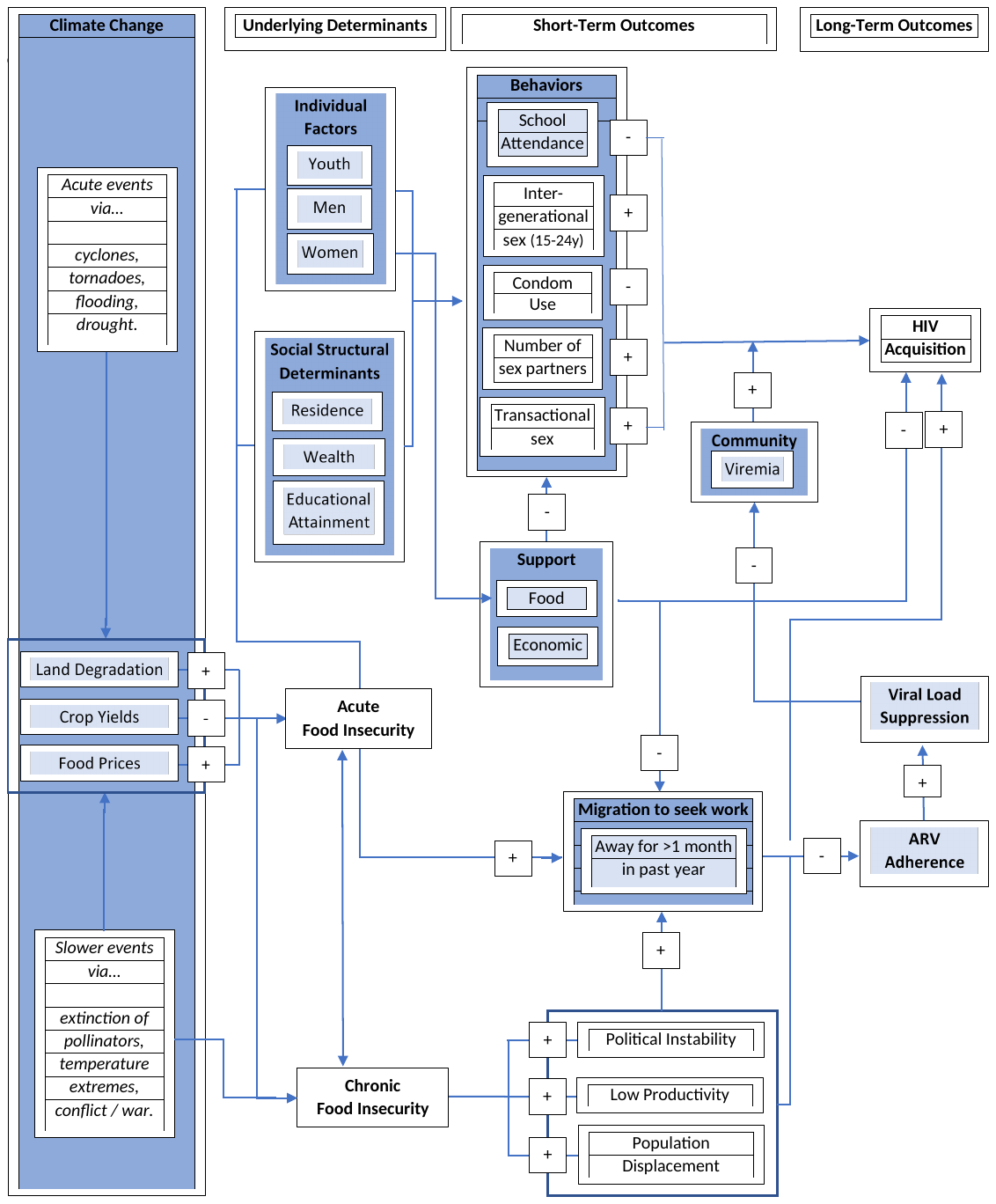

### Climate context preceding and during PHIA data collection

**Global climate context**

Climate extremes have immediate and long-term impacts on livelihoods of poor and vulnerable communities, contributing to greater risks of food insecurity. Different methods of estimating the impact of climate change on food availability have consistently shown that temperature changes will negatively impact crop yields at the global and national levels. With each degree increase in global mean temperature, there would be an average reduction in global yields of wheat by 6%, rice by 3% and soybeans by 3%.^1^ Studies have also shown that in 2019, prior to the COVID-19 pandemic, 34 million people were acutely food insecure. Evidence is also suggesting that 22 million people were displaced due to natural disasters in 2018.^2^ Women comprise the majority of the world’s poor in both the urban and rural sectors and they are the majority of those working in the informal employment sector.^3^

Analysis carried out by the World Food Programme (WFP) on rainfall and temperature patterns in the past 40 years for several countries in the African continent show that while there are marked temperature increases across the region, the case is less clear cut for rainfall. Of the countries under analysis, Uganda, Tanzania and Zambia show positive rainfall trends over the past 40 years, while negative trends are apparent in Eswatini and Lesotho, with no clear trend for Namibia. The trend for Uganda is the most marked (increase of 2.9mm/year) due to an exceptionally wet last three years.

The key rainfall feature for agricultural production and consequently rural food production and food insecurity is the inter-annual (year-on-year) variability in rainfall. In the long run, rainfall variability is a major determinant of livelihoods in the semi-arid tropics as beyond a certain value, purely agriculture-based livelihoods become unfeasible and households switch progressively to livestock-based livelihoods. These fluctuations subject households to the twin hazards of drought and flood. It is a long-term driver of chronic food insecurity as large and unpredictable year-on-year fluctuations in rainfall amounts prevent households from diversifying the crops they plant and lead them to become more risk averse and conservative in terms of their production strategies. In general, the magnitude of inter-annual variations is much larger than any changes arising from a possible long-term trend.

#### *Supplementary Figure 2*. 1981-2020 all country seasonal rainfall for Uganda (left) and Namibia (right). Note large inter-annual fluctuations and lower frequency fluctuations Standardized anomaly of seasonal rainfall for Uganda (left) and Namibia (right) from 1981 to 2020.

**Uganda**	**Namibia**

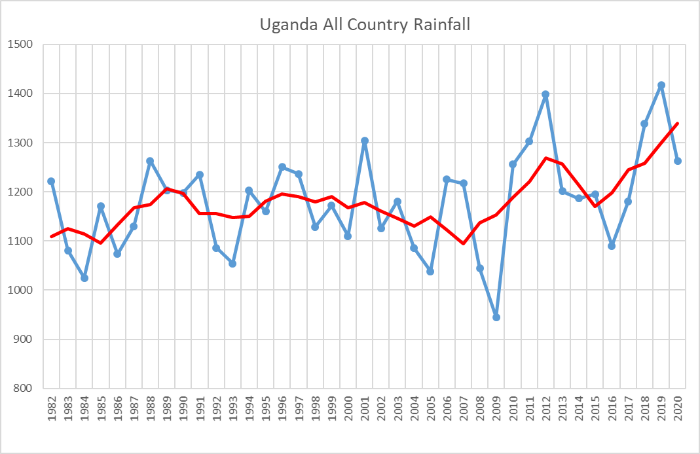

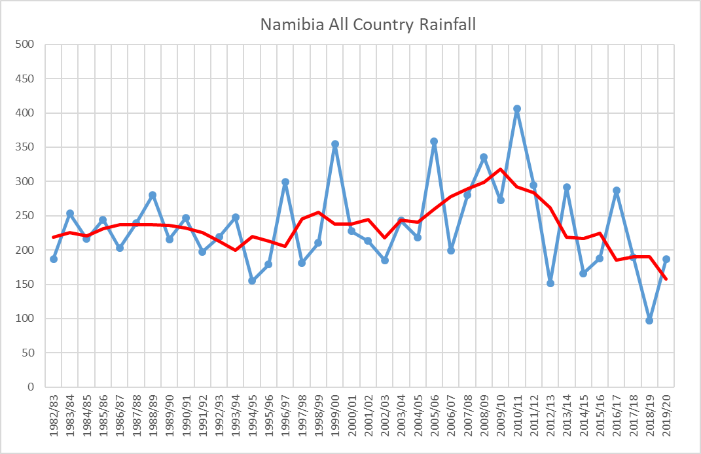

#### *Supplementary Figure 3*. Standardized anomaly of seasonal rainfall for Uganda (left) and Namibia (right) from 1981 to 2020.

**Uganda**	**Namibia**

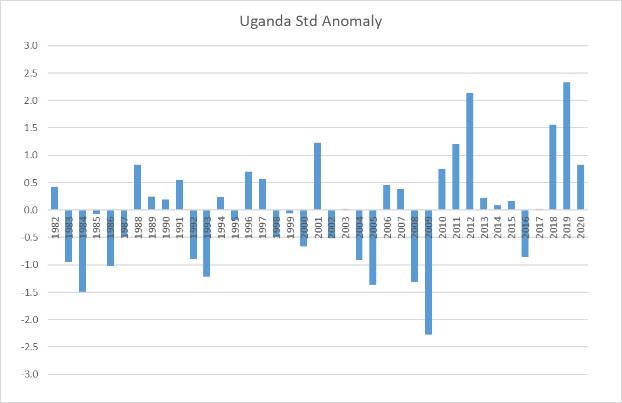

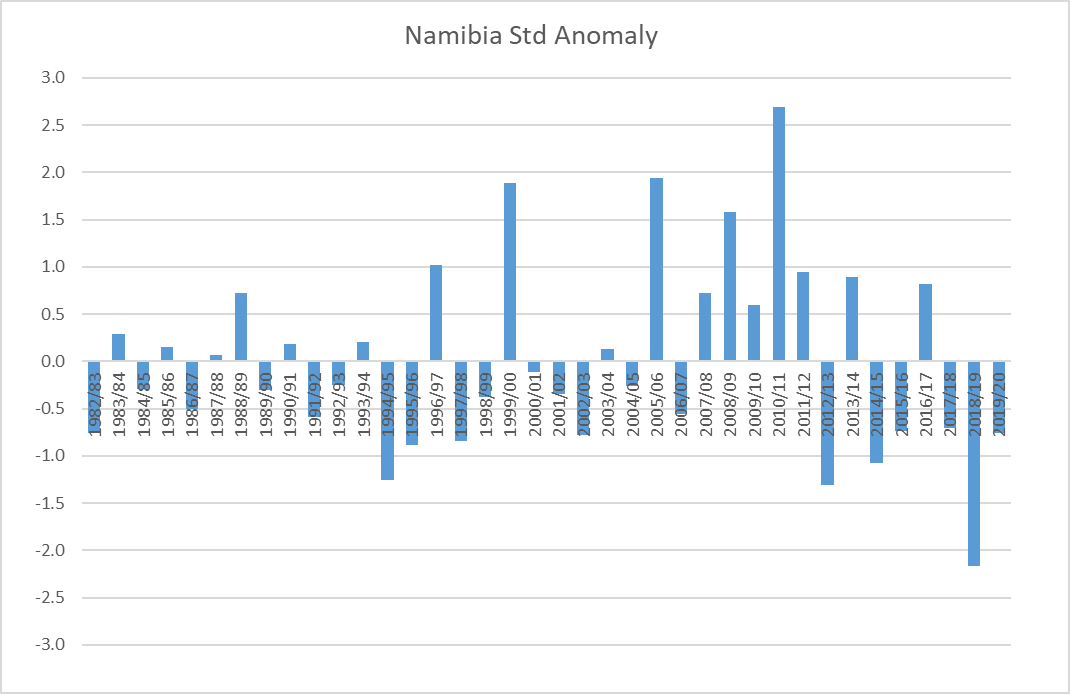

Another mode of variation in seasonal rainfall that may be present is associated with multi-year periods of drier or wetter than average conditions with inter-annual variability super-imposed on these lower frequency cycles. Of the countries in the study, Namibia in particular, but also eSwatini and Lesotho were undergoing drier than average conditions, while both Tanzania and Uganda are in a wetter than average phase.

Two extreme circumstance are exemplified by Uganda and Namibia: since 2010, Uganda has registered a single year with drier than average conditions (2016), while since 2012, Namibia has faced mostly drier than average seasons including the driest year in the 40-year record. Both these situations were preceded by opposite tendencies, a succession of mostly drier than average seasons for Uganda (2002-2009) and wetter than average seasons for Namibia (2005-06 to 2011-12).

The implications are that even if a clear picture were available as to the magnitude and direction of the trend in annual precipitation as a result of climate change, it is the direction and magnitude of change in precipitation variability that would be of crucial importance to infer potential impacts on food insecurity and livelihoods. However, while changes in mean and extreme rainfall have been the object of intense study, rainfall variability has received much less attention. Recent studies indicate that in response to global warming, rainfall variability in tropical areas is expected to increase more than mean precipitation due to greater increase in rainfall extremes.^4^

**Local climate context of PHIA surveys**

It is important to place the timing of the surveys against both the intra-seasonal context and the recent climate context which was somewhat unique at least for Southern Africa. Food insecurity has a well-defined seasonality: typically, in systems dominated by unimodal rain fed agriculture, planting follows soon after the onset of rains with harvests towards the end of the rainfall season. Food insecurity is usually at its lowest after harvest as household stocks are replenished and market prices tend to their yearly minimum. As household stocks are exhausted and staple food prices rise again, food insecurity tends to increase and reach a maximum during the so-called “hunger gap”, a period in the first half of the rainfall season when stocks from the previous harvest have been exhausted, the new harvest is still away and market prices hit their seasonal high. Therefore, following a drought, food insecurity will peak in the early stages of the next rainfall season (even if this happens to be quite favorable).

#### *Supplementary Figure 4.* Timing of Rainfall, Agricultural Cycles, and Food Insecurity

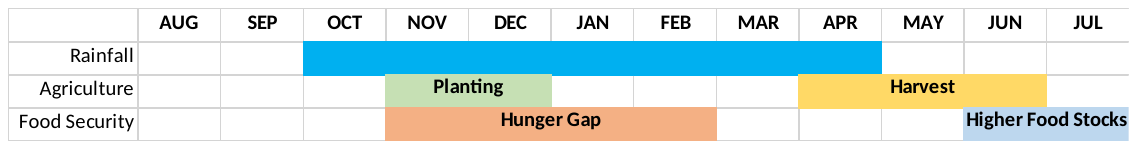

The surveys in this analysis were mostly carried out during a fairly unique period from the climate point of view. From late 2014 to mid-2016 one of the longest and most intense El Niño events on record developed. For Southern Africa in particular, it led to two consecutive droughts, the second of which had very intense impacts on regional food insecurity. Consecutive droughts have compounding effects on food insecurity – the first drought, besides depleting national and regional stocks and direct impacts on households, enhances their vulnerability due erosion of household savings and sale of productive of assets. This enhances the impacts of the second drought through severe reductions in staple food availability and extreme market prices. This was followed by two La Niña events in 2016-2017 and 2017-2018, which nevertheless led to drier than average conditions in East Africa and wetter conditions in Southern Africa.

The figure below shows the timing of the surveys against a simplified drought / food insecurity timeline: we see that the surveys in Zambia took place during harvest time of 2016, after the hunger gap but during a meager harvest and inflated market prices; surveys in eSwatini and Lesotho partly coincided with the period of most extreme food insecurity. In Namibia, the survey took place following the harvests of what was a favorable season, allowing a recovery from the preceding drought impacts. In Tanzania and Uganda, the surveys took place in the drier than average season of 2016-2017. So, except for Namibia, the surveys took place in periods heavily or significantly influenced by drought events.

#### *Supplementary Figure 5*. Climate conditions preceding data collection in each country

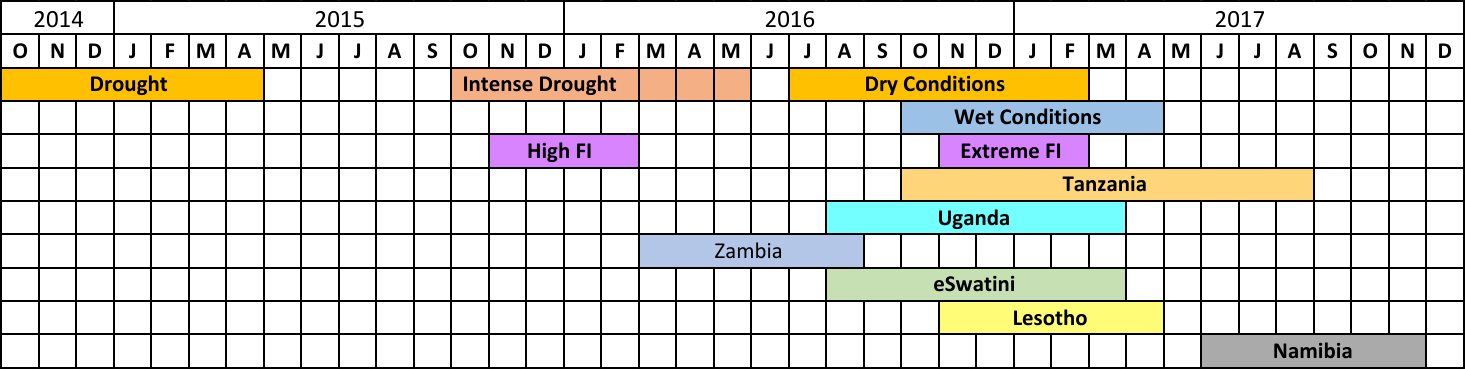

**FI- food insecurity**

### Survey methodology including sample design and construction of variables

**A. Survey Design and Sample Size**

The PHIA surveys employed a cross-sectional, two-stage, cluster sampling design to obtain a nationally representative sample of adults aged 15 years and older, with varying upper age limits.^5-10^ The first-stage sampling units were enumeration areas (EAs) selected with probabilities proportionate to the number of households in the EA, with allocation to subnational areas designed to achieve 30% precision around a national estimate of incidence and 95% confidence intervals (CI) of ±0.10 for regional estimates of viral load suppression (VLS) in individuals aged 15-49 years. There was an assumed intra-cluster correlation of 0^.^05 for prevalence and VLS rates. The estimated numbers of households, individuals and blood draws included adjustments for household vacancy and non-response, number of individuals per household, individual non-response, and refusal of blood testing or specimen loss, based on data derived from the most recent national census. Post-stratification weights were calculated to reflect the age and sex distribution of the most recent national census.

All households within the boundaries of the selected EAs were listed by trained staff prior to data collection. In the second stage of sampling, households were randomly selected from each EA using an equal probability approach that allowed variation in the number of households depending on the size of the EA between the time of the census and the survey household listing. On average, 25 households were selected in each EA.

**B. Variable description**

The food insecurity questions were included in each country’s household questionnaire and included the following questions, based on HFIAS:^11^

In the past 4 weeks, was there ever no food to eat of any kind in your household because of a lack of resources to get food? [YES/NO/DON’T KNOW]

How often did this happen in the past 4 weeks? [RARELY (1-2 TIMES)/SOMETIMES (3-10 TIMES)/OFTEN (MORE THAN 10 TIMES)]?

In the past 4 weeks, did you or any household member go to sleep at night hungry because there was not enough food? [YES/NO.DON’T KNOW]

How often did this happen in the past 4 weeks? [RARELY (1-2 TIMES)/SOMETIMES (3-10 TIMES)/OFTEN (MORE THAN 10 TIMES)]?

In the past four weeks, did you or any household member go a whole day and night without eating anything because there was not enough food? [YES/NO/ DON’T KNOW]

How often did this happen in the past 4 weeks? [RARELY (1-2 TIMES)/SOMETIMES (3-10 TIMES)/OFTEN (MORE THAN 10 TIMES)]?

Heads of households who responded that there was ever no food to eat of any kind their household because of lack of resources to get food, and then classified this as sometime (3-10 times) or often (10 times or more) in the past four weeks, were classified as living in a household with severe food insecurity.

The receipt of economic and food support was asked as:

Has your household received any of the following forms of external economic support in the last 12/3 months?

[SELECT ALL THAT APPLY]

NOTHING [A]

CASH TRANSFER (E.G. PENSIONS, DISABILITY GRANTS, CHILD GRANT) [B]

ASSISTANCE FOR SCHOOL FEES [C]

MATERIAL SUPPORT FOR EDUCATION (E.G. UNIFORMS, SCHOOL BOOKS, EDUCATION, TUITION SUPPORT, BURSARIES) [D]

INCOME GENERATION SUPPORT IN CASH OR KIND (E.G. AGRIGULTURAL INPUTS) [E]

FOOD ASSISTANCE PROVIDED AT THE HOUSEHOLD OR EXTERNAL INSTITUTION [F]

MATERIAL OR FINANCIAL SUPPORT FOR SHELTER [G]

SOCIAL PENSION [H]

OTHER [X]___________________________________(SPECIFY)

DON’T KNOW [Z]

Receipt of food support was defined as having received food assistance provided at the household or external institution in the past three months, regardless of whether they received any other types of support. Other social support included all other types excluding those who received food assistance or who reported having received nothing.

The *socio-demographic characteristics* included residence, defined as urban vs rural, and wealth quintile, which was constructed using Principal Component Analysis (PCA) based on household assets and infrastructure, including the type of house construction, cooking fuel, toilet and water source, based on the methods used by the Demographic and Health Surveys.^12^ For variables at the individual-level, these included age and educational level, defined as the level attended, even if not completed. Employment status was based on reported recent status of paid work, where they were classified as currently enrolled in school, engaged in paid work in the past 12 months, both or neither. Marital status was defined as never married or having lived with a sexual partner, currently married or living with a partner, or no longer married, comprised of all who responded that they were currently separated, divorced or widowed.

*Behavioural variables* included asking whether participants had ever been tested for HIV and received the results, and if they had done so in the past 12 months, and female participants were asked about previous pregnancies and their outcomes. *Sexual behaviour variables* described the lifetime number of sexual partners, who could be partners with whom the participant engaged in either anal or vaginal sex acts. Among those who reported sexual activity in the past 12 months, the following characteristics were measured: how many partners, and for the three most recent partners, their relationship status with the participant (including casual partner, regular partner or husband), their age, and whether they had engaged in the partnership for goods or gifts.

For condom use with an extramarital partner, the denominator was those who reported having an extramarital partner in the past 12 months.

Violence questions were administered to one randomly selected female participant aged 13-59 years in each household. In Tanzania, due to an error in the sampling algorithm, appropriate weights could not be calculated and thus their data was excluded from the analysis.

**Sample weights**

The sample weights were created using similar methodology across all PHIAs.^13^ The sample weights were adjusted to compensate for the variable probabilities of selection for this complex sample design, to account for differential nonresponse rates within relevant subgroups of the sample, and to adjust for under-coverage of certain populations. Taylor weights were used for variance estimates to account for the stratification, clustering and nonresponse and poststratification weighting adjustments. Due to a programming error, violence data were not correctly weighted in Tanzania, and therefore their data were excluded from the analysis of forced sex.

### HIV incidence across the included PHIA countries, by age and sex

#### *Supplementary Figure 6*. HIV Incidence by sex and age among participants across six sub-Saharan countries, 2015-2017

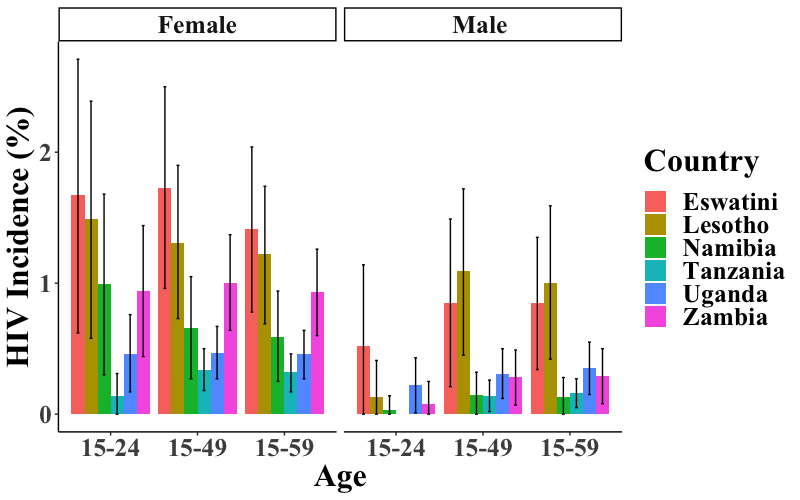

NOTE-Annualized incidence estimates were calculated using the World Health Organization (WHO) incidence formula.^20^

### Measuring food insecurity using PHIA questions to create a hunger scale

Although we used the question on the presence of food in the house to generate our exposure variable, we compared the results to a score generated using all questions. The household hunger scale (HHS) was generated following Ballard et al, (2011).^14^ It uses the six questions listed above, which are collectively validated for identifying household hunger in cross cultural settings. Responses to each of the three questions are scored from 0-2, with zero representing no occurrence of the event (lack of food in the household, going to bed hungry or 24 hours without anything to eat), 1 representing the event occurring “rarely or sometimes”, and 2 representing “often”. The three scores are aggregated to form a continuous household hunger score ranging from 0-6. Then we generated a three-category categorical variable of little hunger, moderate hunger and severe hunger. It is important to note that the continuous household hunger score is generally not normally distributed and therefore use of the mean score for tests of statistical significance is not recommended. The trends in recent infection in women that were observed using the one food in the house variable persisted, but, interestingly, the power was reduced, in large part because the number of participants classified as severely hungry was considerably smaller. This might reflect that the question on whether there was no food in the house is relatively objective, whereas the questions on hunger and going a whole day and night without eating are answered by the head of household for all members of the household, and therefore might not reflect individual household members’ experiences with hunger.

| **Characteristic** | **Women**  **(N=54,784)** | | **Men**  **(N=43,535)** | |
| --- | --- | --- | --- | --- |
|  | **aRR**  **(95% CI)** | **P-value** | **aRR**  **(95% CI)** | **P-value** |
| **Hunger score**  Little  Moderate  Severe | Ref.  1.62 (0.89-2.95)  2.21 (0.38-13.04) | 0.115  0.379 | Ref.  0.66 (0.29-1.50)  3.17 (0.47-21.27) | 0.319  0.234 |
| **Age group**  15-24  25-34  35-44  45-59 | Ref.  1.20 (0.61-2.34)  0.78 (0.35-1.70)  0.35 (0.11-1.15) | 0.598  0.524  0.085 | Ref.  0.84 (0.22-3.17)  1.47 (0.35-6.15)  1.27 (0.29-5.61) | 0.791  0.593  0.755 |
| **Country**  Zambia  Lesotho  Eswatini  Uganda  Namibia  Tanzania | Ref.  0.87 (0.49-1.57)  1.63 (0.88-3.02)  0.69 (0.41-1.14)  0.90 (0.44-1.86)  0.46 (0.26-0.81) | 0.654  0.117  0.147  0.776  0.007 | Ref.  2.26 (0.77-6.67)  2.45 (0.72-8.37)  1.51 (0.59-3.89)  0.62 (0.15-2.55)  0.61 (0.23-1.59) | 0.138  0.153  0.390  0.507  0.310 |
| **Rural residence** | 0.64 (0.36-1.14) | 0.131 | 1.96 (0.80-4.78) | 0.141 |
| **Wealth Quintile** | 0.91 (0.74-1.13) | 0.402 | 0.92 (0.71-1.20) | 0.541 |
| **Community viremia**  **(per 1% increase)** | 1.10 (1.05-1.15) | <0.001 | 1.10 (1.02-1.18) | 0.018 |
| **Receipt of support in past 3 months**  None  Economic only  Food support | Ref.  1.03 (0.54-1.99)  0.36 (0.14-0.90) | 0.921  0.029 | Ref.  1.37 (0.52-3.63)  2.68 (0.37-19.56) | 0.527  0.329 |
| **Marital status**  Never married  Married  Separated/Divorced/Widowed | Ref.  1.87 (0.93-3.75)  4.16 (1.81-9.54) | 0.077  0.001 | Ref.  10.16 (1.93-53.44)  9.48 (1.38-65.18) | 0.006  0.022 |

#### *Supplementary Table 1*. Association between the food insecurity score and recent HIV infection in adults aged 15-59

### Potential mediating behaviors between severe food insecurity and HIV acquisition in women

#### *Supplementary Table 2*. Frequency of mediating behaviors by country, severe food insecurity and age group in women aged 15-59

| **Characteristic** | **Transactional sex**  **% (n/N)** | **Early sexual debut**  **% (n/N)** | **History of forced sex**  **%(n/N)** | **High-risk sex**  **% (n/N)** | **Intergenerational sex in AGYW**  **% (n/N)** |
| --- | --- | --- | --- | --- | --- |
| **Country**  Zambia  Lesotho  Eswatini  Uganda  Namibia  Tanzania | 16.4 (1,201/7,347)  6.1 (304/4,969)  5.0 (194/3,873)  19.5 (2,353/11,824)  9.6 (553/5,908)  13.1 (1,382/11,199) | 9.6 (1,054/10,507)  4.8 (332/6,661)  4.5 (251/5,358)  12.1 (2,009/15,813)  5.8 (586/8,276)  8.7 (1,492/15,884) | 8.1 (496/6,848)  11.9 (503/4,537)  4.1 (98/2,459)  16.2 (384/2,898)  4.5 (211/5,264)  NI | 32.3 (2,395/7,348)  28.7 (1,462/4,976)  22.4 (880/3,869)  38.5 (4,564/11,855)  23.2 (1,593/5,922)  45.8 (5,038/11,246) | 12.3 (257/2,096)  11.3 (161/1,389)  16.3 (160/1,003)  16.3 (630/3,870)  10.2 (182/1,655)  18.2 (616/3,556) |
| **Severe food insecurity**  No  Yes | 14.6 (5,070/39,511)  21.8 (915/5,582) | 9.4 (4,792/54,479)  12.9 (930/7,984) | 7.1 (1,730/26,874)  10.8 (295/3,612) | 39.9 (13,678/39,589)  45.5 (2,243/5,600) | 16.3 (1,744/11,937)  19.0 (261/1,624) |
| **Age group**  15-24  25-34  35-44  45-59 | 19.8 (2,361/13,974)  13.3 (1,821/15,377)  14.3 (1,197/9,586)  10.7 (608/6,183) | 9.5 (2,036/23,227)  9.4 (1,581/17,574)  10.7 (1,140/11,611)  9.9 (967/10,087) | 11.6 (772/7,973)  5.3 (595/9,797)  5.7 (401/6,907)  4.4 (260/5,828) | 37.2 (4,372/14,013)  36.7 (5,007/15,417)  44.6 (3,776/9,607)  53.3 (2,777/6,179) | 16.5 (2,006/13,569)  -  -  - |

Note- analysis restricted to those who report a history of sexual activity aside from sexual debut. History of forced sex did not include data from Tanzania due to sampling error. All percentages are weighted and numbers are crude. Denominators vary due to missing data or different sampling methods for the violence questions. High-risk sex is defined as sex without a condom with someone of unknown or HIV-positive status.

### Severe food insecurity and risk behaviors by country in women aged 15-59

#### *Supplementary Figure 7*. Forest plot of adjusted relative risk for different sexual behaviors in women aged 15-59 by country

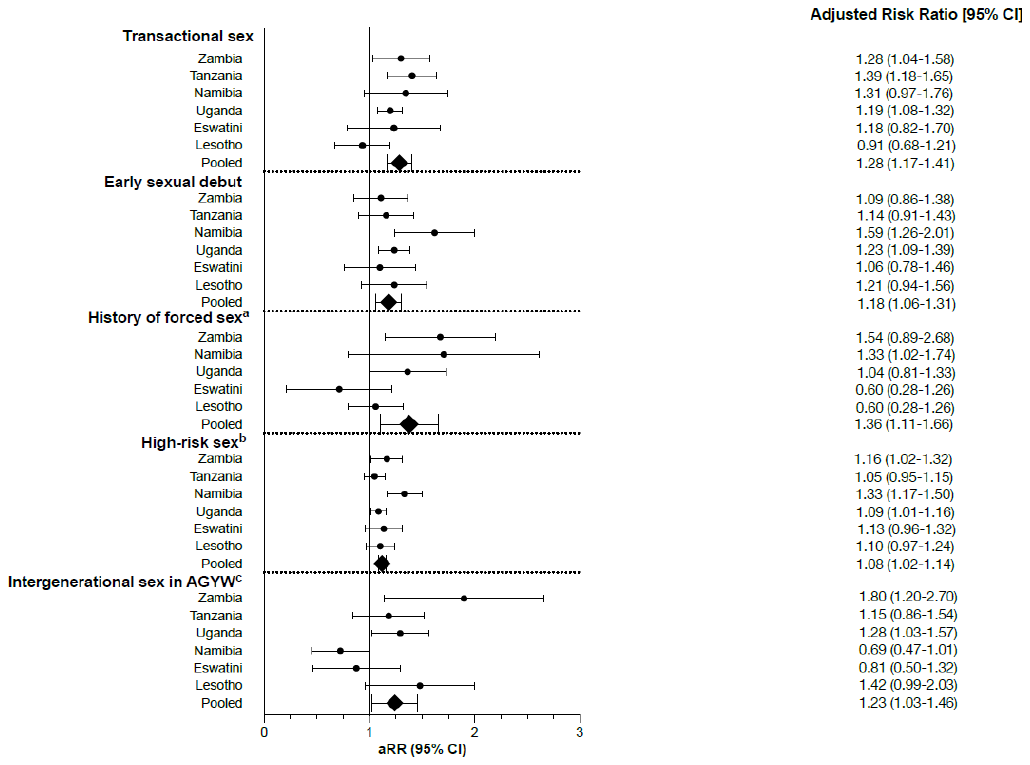

#### aRR- adjusted risk ratio

7.	Ministry of Health Zambia. Zambia Population-based HIV Impact Assessment (ZAMPHIA) 2016: Final report. Lusaka, Zambia, 2019.

8.	Tanzania Commission for AIDS (TACAIDS) ZACZ. Tanzania HIV Impact Survey (THIS) 2016-2017: Final Report. Dar es Salaam, Tanzania, 2018.

9.	Ministry of Health and Social Services (MOHSS) Namibia. Namibia Population-based HIV Impact Assessment (NAMPHIA) 2017: Final Report. Windhoek, Namibia; 2019.

10.	Government of the Kingdom of Eswatini. Swaziland HIV Incidence Measurement Survey 2 (SHIMS2) 2016-2017. Final Report. Mbabane, Eswatini, 2019.

11.	Coates J, Swindale A, Bilinsky P. Household Food Insecurity Access Scale (HFIAS) for Measurement of Household Food Access: Indicator Guide (v. 3). Washington, DC: FHI 360/FANTA; 2007.

12.	Rutstein S. Steps to constructing the new DHS wealth index. 2015.

13.	ICAP at Columbia University, The United States Centers for Disease Control and Prevention, Westat. Population-based HIV Impact Assessment (PHIA) data use manual. July 2019 2019. <http://phia.icap.columbia.edu>.

14.	Ballard T, Coates J, Swindale A, Deitchler M. Household Hunger Scale: indicator definition and measurement guide. In: Food and Nutrition Technical Assistance II Project, editor. Washington, DC,: FHI360; 2011.
